# Mpox severity and mortality in Africa: a systematic review and meta-analysis

**DOI:** 10.1101/2025.11.15.25340267

**Authors:** Fabrice Zobel Lekeumo Cheuyem, Jude Tsafack Zefack, Chabeja Achangwa, Rick Tchamani, Constantine Tanywe Asahngwa

## Abstract

**Background:** Mpox, a zoonotic disease endemic to Africa, presents a significant public health threat with variable severity driven by viral clades and host factors. After over 50 years of outbreaks, this study aimed at providing a comprehensive continent-wide assessment of its severity and mortality for evidence-based public health planning and clinical management.

**Methods:** This systematic review and meta-analysis, conducted per PRISMA guidelines and registered with PROSPERO (CRD420251133745), aggregated data from studies on mpox severity and mortality in Africa from 1970 to 2025. A systematic search of multiple databases was performed. The primary outcomes were the mpox severity rate (proportion of severe cases among confirmed cases) and the case fatality rate (CFR). Random-effects models were used due to high heterogeneity.

**Results:** The analysis included 37 records on severity and 43 on mortality among confirmed cases. The pooled severity rate among confirmed mpox cases in Africa was 43.77% (95% CI: 36.27–51.58). The pooled CFR was 3.92% (95% CI: 1.88–7.60) for confirmed cases and 2.49% (95% CI: 1.32–4.62) for suspected cases. A significant temporal decline was observed; after the 2022 global outbreak, the CFR for confirmed cases dropped to 1.04% from 6.50% beforehand. Substantial geographical disparities existed, with the Central African region bearing the highest burden (severity: 45.51%; CFR: 5.44%), compared to West Africa (severity: 25.43%; CFR: 0.97%). Infections with Clades Ia and Ib were associated with higher severity, whereas Clade II was linked to milder disease. Key risk factors for severe outcomes and death included young age (<10 years), HIV coinfection, lack of prior smallpox vaccination, and pregnancy. Community-based studies reported higher CFRs than hospital-based studies. No significant publication bias was detected, and sensitivity analyses confirmed the robustness of the pooled estimates.

**Conclusion:** This study confirms a high but declining burden of severe and fatal mpox in Africa, characterized by significant temporal, geographic, and clade-specific variations. While a global shift towards milder disease is evident, Central Africa remains disproportionately at risk due to the persistent circulation of more virulent clades and underlying health inequities. Sustained investment in surveillance, vaccination, and strengthened healthcare capacity in endemic regions is crucial to reduce future morbidity and mortality.

## Background

Mpox (formerly known as monkeypox) is a zoonotic disease caused by the monkeypox virus (MPXV), a member of the Orthopoxvirus genus and closely related to the variola virus that caused smallpox. Since its first identification in humans in the Democratic Republic of the Congo (DRC) in 1970, mpox has remained endemic in several African regions, particularly Central and West Africa [1]. Over the last fifty years, the disease has evolved from isolated outbreaks to a pervasive public health concern, characterized by recurring waves of epidemics. Following the end of routine smallpox vaccination in 1980, the population’s cross-protective immunity declined, which paved the way for the reappearance of mpox [2]. Furthermore, its endemicity in Africa has been maintained by growing urbanization, deforestation, wildlife contact, and inadequate healthcare infrastructure.

Although human-to-human transmission of mpox has increased, the disease is essentially a zoonotic one, spread by contact with infected animals such as rodents, primates, and bushmeat. A significant turning point was the global outbreak in 2022, which spread to more than 120 countries and demonstrated the virus’s ability to spread outside endemic areas persistently [3,4]. Recurrent outbreaks have been documented in Africa, including Nigeria, Cameroon, the Democratic Republic of the Congo, and the Central African Republic, underscoring the disease’s tenacity and geographic spread. The World Health Organization recorded 142,151 confirmed cases and 328 deaths worldwide as of May 2025. The global case fatality rate (CFR) was 0.09%, although CFRs in African nations were significantly higher [1].

According to recent genomic studies, the majority of mpox viruses circulating in Africa belong to two distinct clades: Clade I, which has historically been prevalent in Central Africa, and Clade II, which is more common in West Africa [5]. In the DRC, novel sub-lineages of Clade I, such as Clade Ib, have been discovered. They are linked to improved transmissibility but exhibit marginally lower virulence compared to Clade Ia [6]. Clade I infections have traditionally been linked with more severe disease presentations and higher mortality, with CFRs reaching 10% or more in specific outbreaks [7]. Conversely, Clade II, which caused the global 2022–2024 epidemic, has generally been associated with milder disease, lower hospitalization rates, and mortality below 1% [8]. Understanding these clade-specific differences is crucial for informing regionally tailored interventions and clinical management.

Despite mpox’s lengthy history in Africa, there are still few accurate estimates of its severity and fatality rate. Numerous studies currently available are small, geographically limited, and vary in their methods. For instance, hospital-based studies can underrepresent community-level deaths that take place outside of the healthcare system, and surveillance data frequently depend on syndromic or clinical criteria without laboratory validation [9]. Consequently, the actual burden of severe and fatal mpox in Africa remains uncertain, with reported CFRs ranging from below 1% in Nigeria to over 11% in the DRC and Central African Republic [10]. Furthermore, socioeconomic disparities, healthcare access, and vaccination coverage contribute to regional variability. Central Africa continues to report disproportionately higher mortality due to the continued circulation of more virulent Clade I strains and fragile healthcare systems [11].

The severity of mpox infections and their death rate depend on several host-related factors. Studies reported that children under the age of 10 remain the most vulnerable, with the highest rates of hospitalization and fatality [12,13]. Immunocompromised individuals, especially those with advanced HIV infection, are also at increased risk for severe illness due to impaired viral clearance [14]. Similarly, pregnant women are more likely to experience negative consequences, such as miscarriage, fetal death, and problems for the mother [15]. Another important factor influencing the severity of the sickness is the lack of protection from prior smallpox immunization [16]. Malnutrition and co-infection with other tropical diseases, such as chickenpox or malaria, are examples of environmental factors that exacerbate the course and outcome of the disease [17].

The epidemiology of mpox in Africa has undergone a significant transformation between 2022 and 2025, as evidenced by changes in transmission dynamics, as well as a decline in severity and fatality rates. According to surveillance statistics, the percentage of severe cases and fatalities has dropped, even though the total number of confirmed mpox cases has grown [18]. The prevalence of less virulent viral lineages, better clinical therapy, earlier diagnosis, and enhanced case detection are probably the causes of this shift [9]. However, with a pooled CFR of roughly 3–4% compared to less than 0.3% globally, the African continent still has the highest illness burden [1]. These discrepancies highlight ongoing injustices in public health infrastructure, vaccine access, and antiviral availability.

Comprehensive research on mpox outcomes across the continent is desperately needed, given the continuous endemic transmission and frequent outbreaks. This information vacuum can be filled by a systematic review and meta-analysis of the existing literature, which can provide pooled estimates of severity and mortality, examine regional and temporal variations, and identify risk factors associated with unfavorable outcomes. This study aims to generate robust data to inform clinical management recommendations, public health initiatives, and resource allocation by combining over 50 years of research (1970–2024). Furthermore, measuring variations in outcomes among viral clades and health environments can help inform adjustments to response strategies and surveillance priorities in the most affected areas of Africa. By doing this, this study not only sheds light on the historical development of mpox but also helps guide future international readiness initiatives for re-emerging zoonotic illnesses.

## Methods

### Study design

This meta-analysis assesses the epidemiology of the Mpox outbreak in Africa from 1970 to 2024, representing the first such study to evaluate disease severity and mortality on the continent. The review was conducted according to the Preferred Reporting Items for Systematic Review and Meta-analysis (PRISMA) guidelines and is registered with PROSPERO (ID: CRD420251133745) [19,20].

### Eligibility criteria

#### Inclusion criteria

This systematic review and meta-analysis included community- and hospital-based studies (including cohort, cross-sectional, case series, and case report designs) that provided data on mpox severity and/or mortality rates in Africa. Studies were eligible if they reported clinical outcomes or epidemiological surveillance data, such as severe or grave cases, deaths, or case fatality rates. No restrictions were applied on publication year; however, only articles published in English or French were considered.

#### Exclusion criteria

Studies were excluded if they were single case reports, reviews, commentaries without original data, or focused on other viruses or unrelated outcomes (e.g., genetic analyses without clinical correlates). Duplicate publications were removed after title and abstract screening. Studies that lacked clearly defined measures of severity or mortality were also excluded.

### Article searching strategy

A systematic search of electronic databases, including PubMed, Web of Science, Scopus, Embase, CINAHL, and ScienceDirect, was conducted to identify relevant published studies. The search strategy involved querying terms in the titles and abstracts of records. A combination of keywords and Medical Subject Headings (MeSH) was utilized, employing Boolean operators (“AND” and “OR”) to refine the search. The core search string was as follows: (“Mpox” OR “Monkeypox” OR “mpox” OR “monkey pox” OR MPXV) AND (“Disease Severity” OR “severity” OR “severe” OR “complicated” OR “mortality” OR “case fatality” OR “death” OR “died” OR “outcome”) AND (“Africa” OR “Central Africa” OR “West Africa” OR “East Africa” OR “African countries”). To ensure comprehensiveness, a supplementary search was performed using Google Scholar to locate any further relevant publications not indexed in the primary databases. Additionally, the reference lists of all identified studies were screened for other potentially eligible articles. The final search for all sources was completed on June 29, 2025.

### Data extraction

Data were extracted from all eligible articles using a predefined form in Microsoft Excel 2016 to collect study characteristics. The data extraction sheet captured details including the first author’s name, publication year, country, study design, setting, sampling technique, gene sequencing classification, and the number of suspected, confirmed, severe, and fatal mpox cases. Two reviewers (F.Z.L.C. and J.T.Z.) independently assessed each article and extract data. Any discrepancies were resolved through discussion with a third reviewer (C.T.A.) to reach a consensus.

### Data quality assessment

The quality of the included studies was assessed using the Joanna Briggs Institute (JBI) critical appraisal tools [21]. The risk of bias was evaluated using design-specific predefined criteria. Each criterion was scored as “1” (yes) or “0” (no or unclear). The overall risk of bias for each study was then categorized as low (>50% of criteria met), moderate (25-50%), or high (<25%). Two reviewers (F.Z.L.C. and C.A.) independently conducted the assessments, and any disagreements were resolved through consensus discussion.

### Outcome measurement

This study’s primary outcomes were the mpox severity rate and CFR. The severity rate was calculated as the proportion of severe or grave cases among all confirmed mpox cases. The CFR was calculated as the proportion of deaths among populations of suspected or confirmed mpox cases.

### Operational definition

A case was defined as severe or grave if it required hospitalization and presented with one or more of the following: extensive lesions (>100 lesions); hemorrhagic or pustular lesions; mucosal involvement (oral, genital, conjunctival); or systemic complications such as high fever (>39°C), altered general state (e.g., bedridden), sepsis, encephalitis, secondary bacterial infections, hypotension, septic shock, severe dehydration, or keratitis with potential for corneal scarring and blindness [13,22]. A suspected mpox case was defined as an individual presenting with a vesicular or pustular rash—characterized by deep-seated, firm lesions—accompanied by at least one of the following symptoms: fever preceding the rash, lymphadenopathy (inguinal, axillary, or cervical), or lesions on the palms or soles. A case was defined as laboratory-confirmed mpox if at least one specimen yielded a positive result in the rapid Orthopoxvirus-specific assay, or Mpox-specific real-time PCR, or mpox in culture [13].

### Statistical analysis and synthesis

Heterogeneity between studies was quantified using the *I*^*2*^ statistic and categorized as low (<25%), moderate (25–75%), or high (>75%). Given the anticipated high heterogeneity, a random-effects model was employed for all meta-analyses. To explore potential sources of heterogeneity, we conducted subgroup analyses based on study period (defined as pre- and post-2022 global outbreak), country, WHO African region, clade, study design, and setting. Countries were categorized into WHO African regions as follows: Western (Sierra Leone, Liberia, Nigeria), Eastern (Burundi, Sudan), and Central (Cameroon, Central African Republic, Gabon, Congo, and the DRC). Furthermore, meta-regression was performed to assess the association between these study characteristics and the variability in effect sizes [23].

Univariable and multivariable meta-regression models were employed to evaluate the association between study-level covariates and the pooled severity and CFR. We used Generalized Linear Mixed Models (GLMM) with a Probit-Logit Transformation (PLOGIT), which are recommended for synthesizing binomial proportion data as they directly model the binomial distribution, incorporate random effects to account for between-study heterogeneity, and naturally handle proportions of 0% and 100% without ad-hoc correction[24]. A two-sided *p*-value < 0.05 was considered statistically significant. All analyses were conducted in R Statistics version 4.4.2 using the “meta” package [25].

### Publication bias and sensitivity test

Publication bias was assessed both visually and statistically. Funnel plots were generated for visual inspection, where symmetry suggests a low risk of bias. Statistical evaluation was performed using Egger’s linear regression test and Begg’s rank correlation test, with a *p*-value < 0.05 indicating potential publication bias. To assess the robustness of the pooled estimates, a leave-one-out sensitivity analysis was conducted by iteratively removing each study and recalculating the summary effect.

## Results

A total of 5443 records were retrieved from the database search (*n* = 5438) and from other sources (*n* = 5). After removing 732 duplicate records, 4706 records remained. Titles/abstracts, followed by full text articles were then screened for eligibility. Ultimately, 19 study reports met the eligibility criteria and were included in the meta-analysis (Fig. 1).

**Fig. 1.**
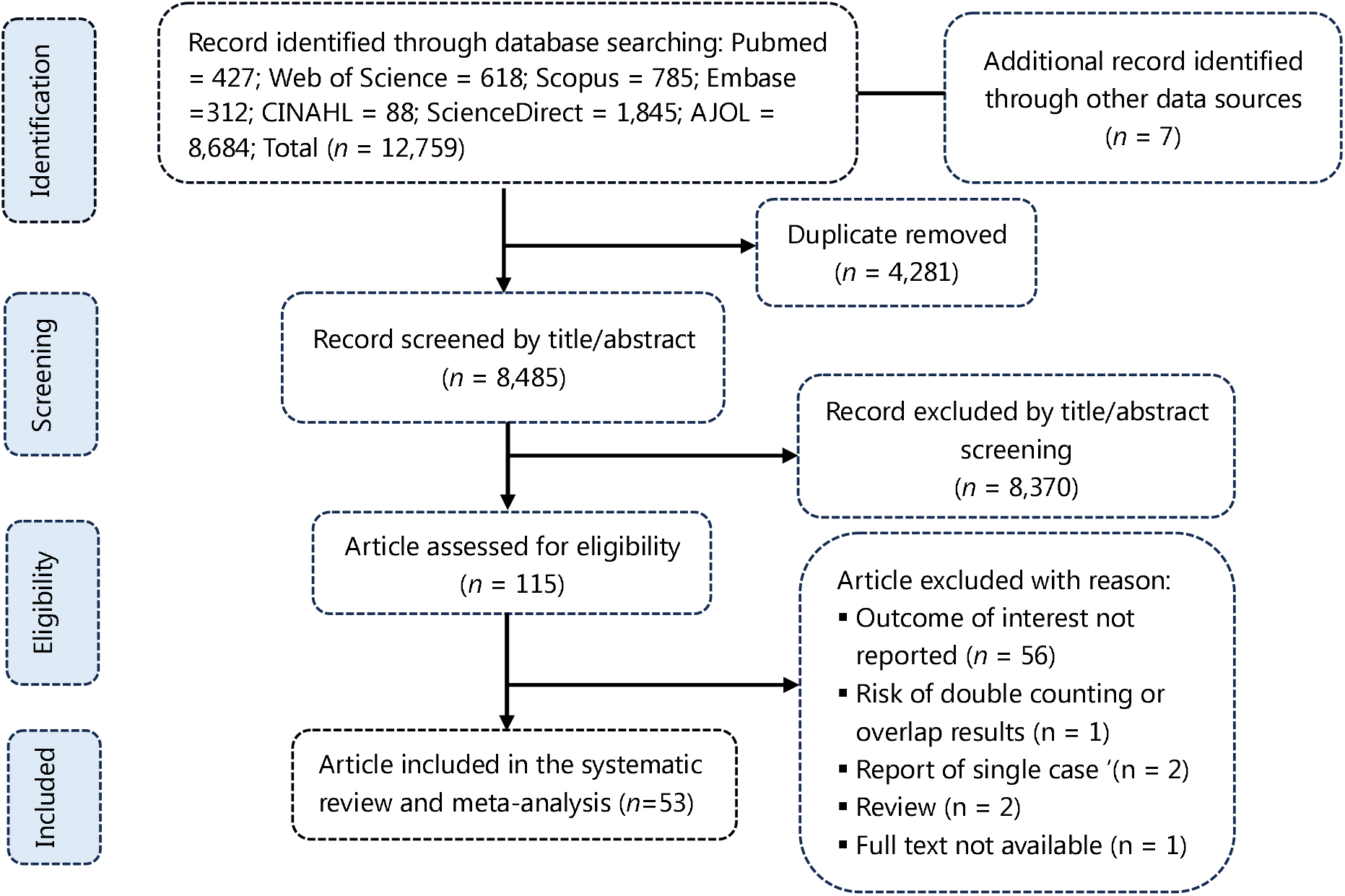
PRISMA diagram flow from study identification to inclusion in the meta-analysis

### Studies selection

### Characteristic of reports included

This dataset aggregated evidence from 37 records across 10 African countries, documenting mpox severity from 1970 to 2024 [6,7,11,13,15,17,26–54]. The DRC was the most frequently studied country (n = 15 records). The majority of reports (n = 28 records) were cross-sectional in design and conducted in community settings (n = 27 records). Gene sequencing data, reported in 12 studies, indicates the presence of Clades Ia, Ib, and II, with Clade Ib being the most commonly reported in recent years (2022-2024). The risk of bias was predominantly low (n = 33 records) (Additional file 1, Supplementary Table 1).

The second outcome derived from 43 records across 11 African countries, documenting mpox case fatality from 1970 to 2024 [7,11,12,15,26–33,36–43,45–48,50– 53,55–59]. The DRC and the Central African Republic (CAR) are the most frequently studied countries. Study designs were predominantly cross-sectional (n = 30 records) and conducted in community settings (n=28 records). Gene sequencing data, reported in 11 records, indicates the presence of Clades Ia, Ib, and II. The risk of bias was mostly low (n = 32 records) or moderate (n = 11 records) (Additional file 2, Supplementary Table 1).

The third outcome resulted from 47 records on suspected mpox cases across 11 African countries, spanning 1970 to 2024 [6,7,15,16,30–33,36–41,41–43,46– 48,50,52,53,55–58,58–65]. The DRC, Nigeria, and the Central African Republic (CAR) are the most frequently studied regions. Study designs are predominantly cross-sectional (n=31 records) and were conducted in both community (n=28 records) and hospital (n=10 records) settings. The risk of bias was primarily low (n=30 records) or moderate (n=15 records) (Additional file 3, Supplementary Table 1).

### Disease severity among confirmed mpox cases

This study found that the pooled disease severity rate for mpox in Africa was 43.77% (95% CI: 36.27–51.58), based on 37 records. The analysis demonstrated high heterogeneity among the included studies (*I*^2^ = 87.5%) (Fig. 2).

**Fig. 2.**
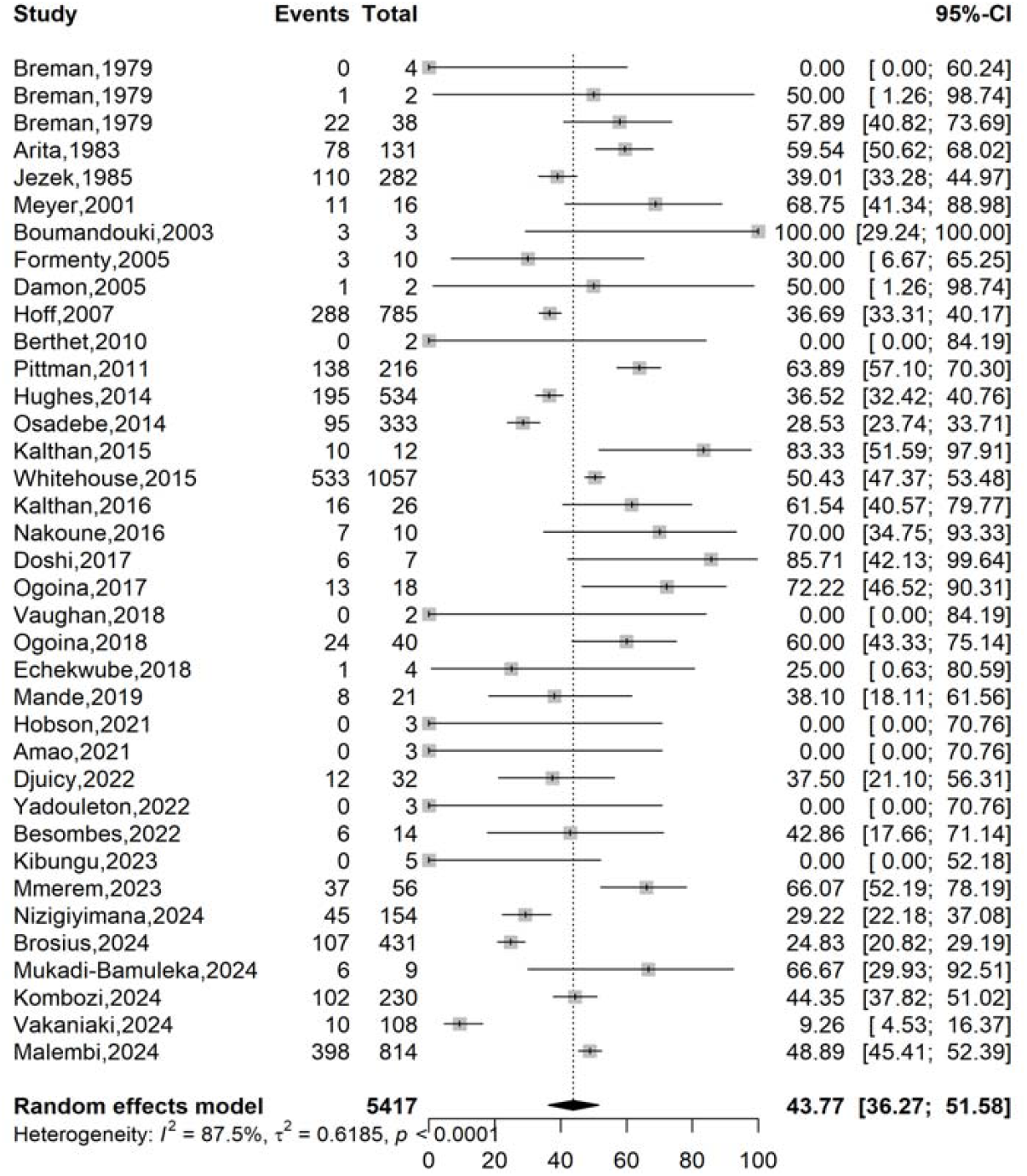
Pooled severity rate of Mpox case in Africa

A non-significant decrease in severity was observed between the period before the 2022 global outbreak (49.22%; 95% CI: 41.44–57.04%; *n* = 26 records) and the period after (34.11%; 95% CI: 22.61–47.84%; *n* = 11 records). Furthermore, a significant difference in severity was found among confirmed mpox cases across WHO African regions [p(*χ*^2^) < 0.001]. Countries in the Central African region exhibited the highest pooled proportion of severe cases (45.51%; 95% CI: 36.55–54.77%; *n* = 23 records), while the lowest proportion was observed in West African countries (25.43%; 95% CI: 6.49– 62.64%; *n* = 10 records). The most affected countries included Congo (90%; 95% CI: 53.28–98.61%; *n* = 2 records) and the Central African Republic (60.92%; 95% CI: 47.54– 72.83%; *n* = 5 records) (Fig. 3).

**Fig. 3.**
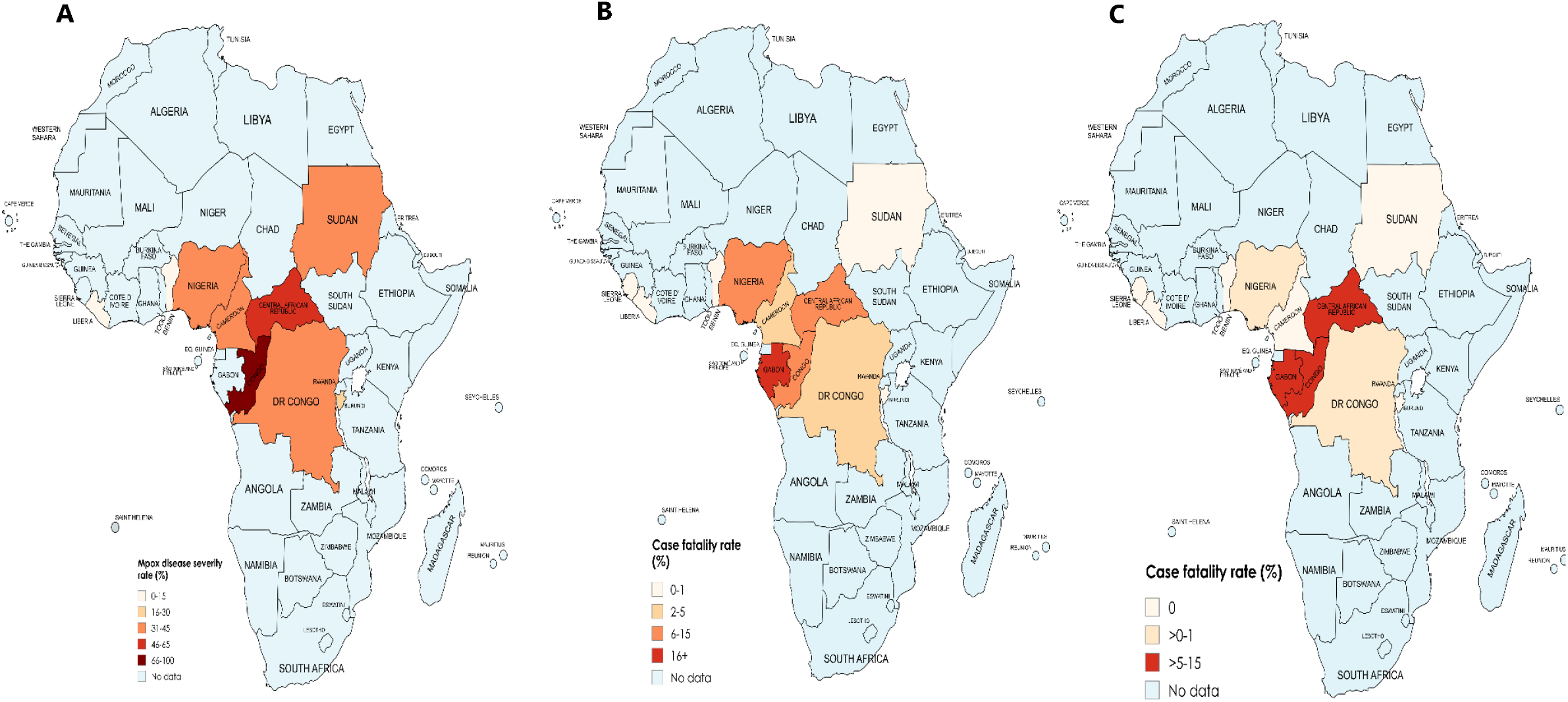
Mapping of mpox severity (A) and case fatality rates among confirmed (B) and suspected (C) mpox cases across affected African

Stratification by gene sequencing showed that cases infected with Clades Ia and Ib had a significantly higher severity rate (30.30%; 95% CI: 7.15–47.74%; *n* = 5 records) compared to those infected with Clade II (0%; *n* = 3 records) (Table 1 and Additional File 1, Supplementary Fig. 1-6).

**Table 1.** Subgroup meta-analysis of mpox severity rate pooled estimates in Africa.

| Subgroup | Confirmed cases | Event rate<sup>1</sup> (%) | 95% CI limits<sup>1</sup> |  | Number of studies | Heterogeneity statistic<sup>1</sup> |  |
| --- | --- | --- | --- | --- | --- | --- | --- |
|  |  |  | Lower | Upper |  | *I*<sup>2</sup>(%) | *p*-value |
| **Study period<sup>2</sup>** |  |  |  |  |  |  |  |
| <2022 | 3561 | 49.22 | 41.44 | 57.04 | 26 | 84.2 | <0.001 |
| 2022-2024 | 1856 | 34.11 | 22.61 | 47.84 | 11 | 91.9 | <0.001 |
| **Region<sup>3</sup>** |  |  |  |  |  |  |  |
| Eastern | 166 | **29.52** | 23.08 | 36.89 | 3 | 0.0 | - |
| Central | 4985 | **45.51** | 36.55 | 54.77 | 23 | 90.8 | <0.001 |
| Western | 135 | **25.43** | 6.49 | 62.64 | 10 | 0.0 | - |
| Multi-region | 78 | **59.54** | 50.62 | 68.02 | 1 | - | - |
| **Clade** |  |  |  |  |  |  |  |
| Ia | 33 | 30.30 | 17.15 | 47.74 | 5 | 0.0 | - |
| Ib | 1516 | 30.75 | 16.56 | 49.84 | 5 | 96.3 | <0.001 |
| II | 8 | 0.00 | 0.00 | 100.00 | 3 | 0.0 | - |
| I & II | 32 | 37.50 | 21.10 | 56.31 | 1 | - | - |
| Not reported | 3828 | 52.10 | 44.73 | 59.39 | 24 | 86.9 | <0.001 |
| **Setting** |  |  |  |  |  |  |  |
| Community | 3798 | 41.47 | 32.67 | 50.85 | 26 | 84.4 | <0.001 |
| Hospital | 773 | 50.79 | 33.77 | 67.63 | 9 | 92.9 | <0.001 |
| Community and hospital | 846 | 43.77 | 36.27 | 51.58 | 2 | 36.5 | 0.210 |
| **Design** |  |  |  |  |  |  |  |
| Cohort | 487 | 43.68 | 18.34 | 72.80 | 2 | 97.1 | <0.001 |
| Cross-sectional | 4899 | 45.61 | 38.05 | 53.38 | 26 | 88.4 | <0.001 |
| Cases serie | 22 | 30.72 | 1.37 | 93.41 | 5 | 0.0 | - |
| Case report | 9 | 11.11 | 1.54 | 49.99 | 4 | 0.0 | - |

<sup>1</sup>Random effects model; CI: Confidence Interval; <sup>2</sup>Period before and after the global outbreak; <sup>3</sup>Eastern: Burundi and Sudan; Central: Cameroon, Central African Republic, Congo, and the Democratic Republic of Congo; Western: Benin, Liberia, and Nigeria; Bold: significant difference between tested subgroups ( $p(\chi^2) < 0.05$ ).

### Disease mortality among confirmed mpox cases

A confirmed mpox case had a pooled probability of 3.92% (95% CI: 1.88–7.60; *n* = 43 records) to die because of the disease with high heterogeneity between studies (*I*^2^ = 84.2) (Fig 4). A significant reduction in the case fatality rate was observed between the period before (6.50%; 95% CI: 3.35–12.23; *n* = 35 records) and after the 2022 global mpox outbreak (1.04%; 95% CI: 0.30–3.51; *n* = 8 records) [*p*(*χ*^2^) = 0.009].

**Fig. 4.**
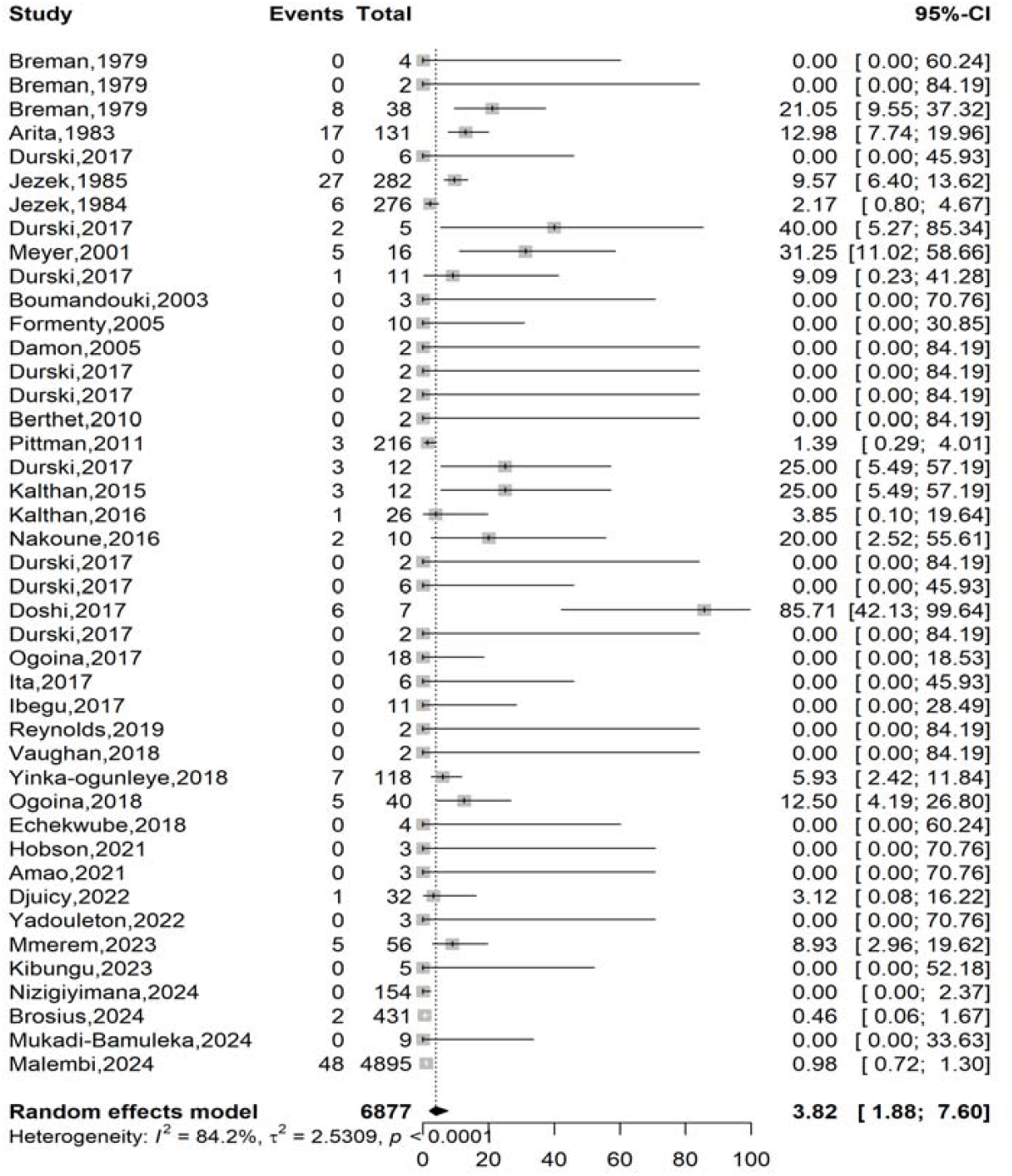
Pooled case fatality rate among confirmed mpox case in Africa

**Fig. 5.**
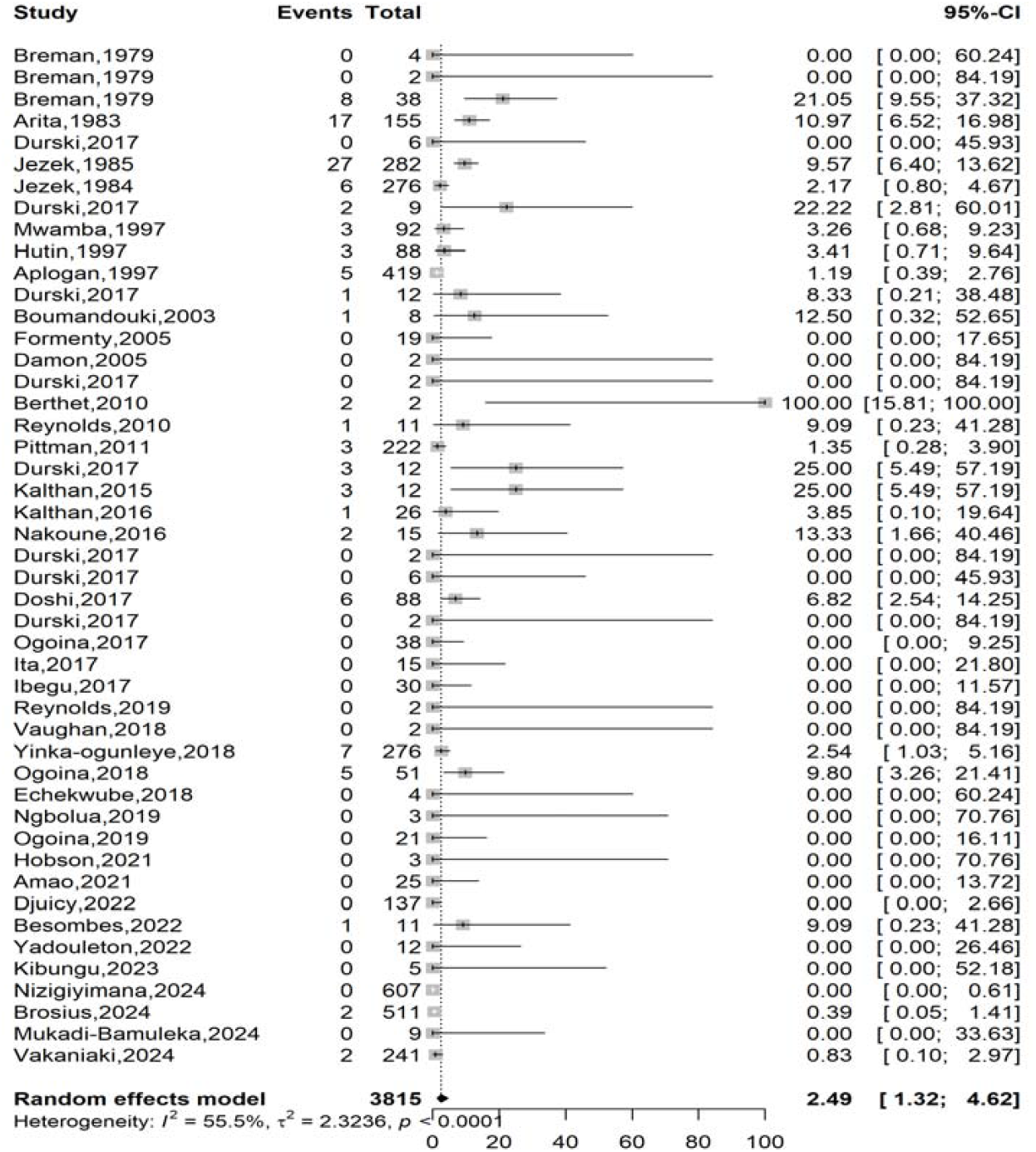
Pooled case fatality rate among suspected Mpox case in Africa

Studies from community settings reported a significantly higher case fatality rate (6.11%; 95% CI: 2.68–13.20; n = 30 records) than those conducted in hospital settings (1.77%; 95% CI: 0.49–6.21; *n* = 11 records) [*p*(*χ*^2^) < 0.001].

Although not statistically significant, Clade II demonstrated a higher case fatality rate (5.56%; 95% CI: 2.67–11.20; *n* = 4 records) compared to Clade Ia (0.00%; *n* = 4 records) and Clade Ib (0.91%; 95% CI: 0.20–4.07; n = 5 records). Similarly, the WHO African regions of Western (6.20%; 95% CI: 3.89–9.75; *n* = 15 records) and Central Africa (5.44%; 95% CI: 2.37–12.01; *n* = 24 records) had higher mortality rates than the Eastern African region (0.00%; n = 3 records) (Table 2, Fig 3, Additional file 2, Supplementary Figs. 1–6).

**Table 2.** Subgroup meta-analysis of case fatality rate pooled estimates for confirmed mpox cases in Africa.

| Subgroup | Confirmed cases | Event rate<sup>1</sup> (%) | 95% CI limits<sup>1</sup> |  | Number of studies | Heterogeneity statistic<sup>1</sup> |  |
| --- | --- | --- | --- | --- | --- | --- | --- |
|  |  |  | Lower | Upper |  | I<sup>2</sup>(%) | p-value |
| Study period<sup>2</sup> |  |  |  |  |  |  |  |
| <2022 | 1292 | 6.50 | 3.35 | 12.23 | 35 | 49.8 | <0.001 |
| 2022-2024 | 5585 | 1.04 | 0.30 | 3.51 | 8 | 71.5 | <0.001 |
| Region<sup>3</sup> |  |  |  |  |  |  |  |
| Eastern | 166 | 0.00 | 0.00 | 100.00 | 3 | 0.0 | 1.000 |
| Central | 6306 | 5.44 | 2.37 | 12.01 | 24 | 90.0 | <0.001 |
| Western | 274 | 6.20 | 3.89 | 9.75 | 15 | 0.0 | 1.000 |
| Multi-region | 131 | 12.98 | 7.74 | 19.96 | 1 | - | - |
| Clade |  |  |  |  |  |  |  |
| Ia | 19 | 0.00 | 0.00 | 100.00 | 4 | 0.0 | 1.000 |
| Ib | 5545 | 0.91 | 0.20 | 4.07 | 5 | 83.0 | <0.001 |
| II | 126 | 5.56 | 2.67 | 11.20 | 4 | 0.0 | 1.000 |
| I & II | 32 | 3.12 | 0.08 | 16.22 | 1 | - | - |
| Not reported | 1155 | 7.25 | 3.61 | 14.03 | 30 | 56.8 | <0.001 |
| Setting |  |  |  |  |  |  |  |
| Community | 1160 | 6.11 | 2.68 | 13.30 | 30 | 45.9 | 0.004 |
| Hospital | 790 | 1.77 | 0.49 | 6.21 | 11 | 56.1 | 0.012 |
| Community and hospital | 4927 | 0.99 | 0.75 | 1.31 | 2 | 24.5 | 0.250 |
| Design |  |  |  |  |  |  |  |
| Cohort | 487 | 3.82 | 1.88 | 7.60 | 2 | 92.2 | <0.001 |
| Cross-sectional | 6357 | 4.51 | 2.38 | 8.38 | 31 | 87.1 | <0.001 |
| Cases serie | 24 | 0.00 | 0.00 | 100.00 | 6 | 0.0 | 1.000 |
| Case report | 9 | 0.00 | 0.00 | 100.00 | 4 | 0.0 | 1.000 |

<sup>1</sup>Random effects model; CI: Confidence Interval; <sup>2</sup>Period before and after the global outbreak; <sup>3</sup>Eastern: Burundi and Sudan; Central: Cameroon, Central African Republic, Congo, and the Democratic Republic of Congo; Western: Benin, Liberia, and Nigeria; Bold: significant difference between tested subgroups ( $p(\chi^2) < 0.05$ ).

### Disease mortality among suspected mpox cases

Suspected mpox case fatality rate was 2.49% (95% CI: 1.32-4.62) based on 47 records (Fig. 6). The subgroup analysis revealed significant temporal and geographical disparities in mpox severity. The case fatality rate was significantly higher in the period before the 2022 global outbreak (4.33%, 95% CI: 2.65–7.01; n = 39 records) compared to the period after (0.31%, 95% CI: 0.08–1.14; n = 8 records) [*p*(*χ*^2^) < 0.001]. Geographically, the Central African region significantly bore the highest burden with a fatality rate of 4.04% (95% CI: 2.15–7.45; n = 28 records), contrasting with the lower rates in Western Africa (0.97%, 95% CI: 0.08–10.33; n = 15 records) [*p*(*χ*^2^) < 0.025] (Table 3, Fig. 3 and Additional file 3, Supplementary figure 1-5).

**Table 3.** Subgroup meta-analysis of case fatality rate pooled estimates among suspected mpox cases in Africa.

| Subgroup | Confirmed cases | Event rate<sup>1</sup> (%) | 95% CI limits<sup>1</sup> |  | Number of studies | Heterogeneity statistic<sup>1</sup> |  |
| --- | --- | --- | --- | --- | --- | --- | --- |
|  |  |  | Lower | Upper |  | I<sup>2</sup>(%) | p-value |
| Study period<sup>2</sup> |  |  |  |  |  |  |  |
| <2022 | 2282 | 4.33 | 2.65 | 7.01 | 39 | 50.5 | <0.001 |
| 2022-2024 | 1533 | 0.31 | 0.08 | 1.14 | 8 | 0.0 | 0.470 |
| Region<sup>3</sup> |  |  |  |  |  |  |  |
| Eastern | 628 | 0.00 | 0.00 | 100.00 | 3 | 0.0 | 1.000 |
| Central | 2545 | 4.04 | 2.15 | 7.45 | 28 | 70.2 | <0.001 |
| Western | 487 | 0.97 | 0.08 | 10.33 | 15 | 0.0 | 0.977 |
| Multi-region | 155 | 10.97 | 6.52 | 16.98 | 1 | - | - |
| Setting |  |  |  |  |  |  |  |
| Community | 2770 | 3.71 | 1.97 | 6.90 | 34 | 57.1 | <0.001 |
| Hospital | 908 | 1.06 | 0.28 | 4.00 | 12 | 43.9 | 0.051 |
| Community and hospital | 137 | 0.00 | 1.32 | 4.62 | 1 | - | - |
| Design |  |  |  |  |  |  |  |
| Cohort | 511 | 0.39 | 0.05 | 1.41 | 1 | - | - |
| Cross-sectional | 3167 | 2.40 | 1.22 | 4.66 | 36 | 59.3 | <0.001 |
| Cases serie | 128 | 6.25 | 3.16 | 12.00 | 6 | 0.0 | 0.999 |
| Case report | 9 | 0.00 | 0.00 | 96.31 | 4 | 0.0 | 1.000 |

<sup>1</sup>Random effects model; CI: Confidence Interval; <sup>2</sup>Period before and after the global outbreak; <sup>3</sup>Eastern: Burundi and Sudan; Central: Cameroon, Central African Republic, Congo, and the Democratic Republic of Congo; Western: Benin, Liberia, and Nigeria; Bold: significant difference between tested subgroups ( $p(\chi^2) < 0.05$ ).

### Meta-regression analysis

According to multivariate meta-regression, no study-related factors significantly influenced the pooled severity rate. However, for case fatality, study design and setting were significant predictors. Case series designs were associated with higher fatality rates (*β* = 1.7165; *p* = 0.044), while in-hospital studies were associated with lower rates (*β* = -1.6932; *p* = 0.007). Among suspected cases, case reports predicted higher fatality (*β* = 2.5908; *p* = 0.007) (Table 4).

**Table 4** Univariate and multivariate meta-regression analysis of Mpox disease outcome metrics in Africa

### Publication bias and sensitivity test analysis

No evidence of publication bias or undue influence from any single study was found for the mpox severity rate (Additional file 1, Supplementary 7 and 8).

Using the trim and fill method to account for 18 potentially unpublished studies yielded a case fatality rate of 3.27% (95% CI: 2.01-5.58) among confirmed mpox cases, which was not significantly different from the initial estimate of 3.92% (95% CI: 1.88–7.60) derived from 43 records. Sensitivity analysis confirmed that while the estimate from one study (Malembi *et al*., 2024) [11] was influential, the overall pooled case fatality rate remained robust and was not disproportionately affected by any single study (Additional file 2, Supplementary Fig. 7 and 8).

For the pooled case fatality rate among suspected mpox cases, the trim and fill analysis did not impute any missing studies, despite a significant sign of publication bias indicated by Begg’s test (*p* = 0.019). Furthermore, sensitivity analysis showed that no single study significantly influenced the pooled estimate, reflecting the robustness of this finding (Additional file 3, Supplementary Fig. 6 and 7).

### Review of severity and mortality-related factors

#### Age

Children, particularly those under 10 years of age, have been identified as an at risk population for mpox, associated with a more severe disease prognosis and a higher mortality rate [12,13,16,26,28,35,55,60]. Vulnerability is highest among young children aged 0-4 years, who consistently present an increased risk of severe complications, including death [6,12,13,26,28]. In adults, severe forms of the disease and mortality are frequently observed in individuals of reproductive age, particularly in cases of HIV co-infection, which is a major factor in worsening the prognosis [7,40].

#### HIV coinfection

Coinfection with HIV, especially in individuals with advanced immunosuppression (low CD4 counts), is a strongly risk factor for both severe mpox manifesting as extensive and prolonged skin lesions, higher complication rates, and significantly increased mortality [6,7,15,40,43,50].

#### Viral clade

Infection with Clade I (Congo Basin) is a major demographic and virological risk factor associated with more severe clinical disease, including higher lesion counts and complication rates, and a significantly higher case fatality rate (CFR) historically up to 10% or more, compared to the milder disease and lower mortality (<1% CFR) associated with Clade II [11,31,47,50].

#### Vaccination status

The absence of cross-protective immunity from prior smallpox vaccination is associated with more severe mpox disease, a higher rate of sequelae, and a significantly increased risk of mortality compared to vaccinated individuals [12,16,26,28].

#### Pregnancy

Mpox infection during pregnancy is a risk factor for severe maternal morbidity and poses a high risk of vertical transmission, placental infection, fetal demise, and in some cases, maternal mortality [11,15].

#### Route of exposure and inoculum

Severe cases were often linked to severe initial exposure, such as from hunting, skinning, or preparing bushmeat from infected animals, suggesting a high inoculating viral dose [12,27,28].

#### Viraemia

Detection of the virus in the blood indicates systemic infection and is associated with severe disease [11].

#### Sexual transmission and genital lesions

Genital lesions, which constitute both a severity factor and a transmission vector for mpox, facilitate the virus’s sexual spread [6,11].

#### Coinfections with other pathogens and sepsis

Concurrent infections (e.g., with Varicella-Zoster Virus or malaria) can complicate the clinical picture, delay diagnosis, and exacerbate systemic illness, leading to severe disease [17,34,51]. Secondary bacterial infections of skin lesions are a common pathway to sepsis, a frequent cause of death [12,17,34].

#### Hemorrhagic manifestations

The development of hemorrhagic symptoms is a rare but grave prognostic sign indicative of severe systemic involvement and coagulopathy, and it is often associated with a fatal outcome [12].

#### Underlying immunosuppressive conditions and malnutrition

Conditions such as malnutrition, untreated tuberculosis, or iatrogenic immunosuppression impair the host response and are common background factors associated with severe disease, particularly in pediatric cases, and contribute to poorer outcomes [13,36].

#### Delay in presentation and critical organ involvement

Patients presenting late in the disease course, or with evidence of critical complications such as encephalitis, bronchopneumonia, or extensive organ involvement, have a markedly poorer prognosis and higher risk of mortality [40,59].

Nosocomial transmission and hospitalization

Outbreaks in healthcare settings can expose vulnerable populations (e.g., malnourished children, immunocompromised individuals). The reason for hospitalization is often itself an indicator of severity [38].

#### Health status

Poor health status of the population was a contributing factor to the high mortality observed [29].

## Discussion

This meta-analysis provides a comprehensive assessment of mpox severity and mortality in Africa from 1970 to 2024. The pooled estimates indicate that 43.8% of confirmed mpox cases were severe, with CFR of 3.9% among confirmed cases and 2.5% among suspected cases. These findings confirm that mpox remains a significant public health threat, with a substantial burden concentrated in the Central Africa region where the disease is endemic. However, a declining trend in both severity and mortality in recent years suggests positive developments, potentially due to improved disease recognition, enhanced clinical management, and the increased prevalence of less virulent viral clades.

### Temporal trends in mpox severity and Mortality in Africa

This meta-analysis demonstrates a clear temporal decline in disease severity. Before the 2022 global outbreak, severity was 49.22%, declining to 34.11% after 2022, while the confirmed case CFR (Case Fatality Rate) dropped from 6.50% to 1.04%. These results are consistent with earlier literature, which reported higher fatality during the 1980s and 1990s in the central African region [12].

In contrast, our post-2022 estimate closely aligns with global surveillance data showing a CFR below 0.3%, based on more than 114,000 cases across more than 120 countries during 2022–2024 [1,8]. The reduction of cases globally coincides with the predominance of Clade Ib, which is linked to milder clinical presentation and low hospitalization rates in non-endemic regions [3]. Yet, Africa’s pooled CFR of 3.9% remains markedly higher than global averages (< 0.3%), underscoring persistent disparities in viral clade distribution, healthcare access, surveillance capacity, and poor vaccination coverage across the African continent [8].

However, recent data from the WHO as of 2024 indicate a substantial decline in CFRs, with contemporary estimates of approximately 0.5% in 2024 across endemic African regions. This improvement likely reflects enhanced diagnostic capacity, expanded surveillance (with >95% of suspected cases laboratory-confirmed in most countries), earlier clinical management, and the gradual shift in circulation toward Clade Ib mpox virus, which appears less virulent than the historical Clade I lineage [1].

Importantly, recent field investigations in the DRC reveal outbreaks dominated by Clades I and Ib, associated with increased transmissibility and severe outcomes among children [9]. These findings confirm that while mpox severity and mortality are declining globally, endemic African regions remain at disproportionately high risk due to persistent Clade I circulation and limited immunization coverage.

### Regional and demographic disparities

Our review also identified substantial geographical heterogeneity. The Central African region had the highest severity (45.51%) and CFR (5.44%), compared to West Africa (25.43% severity; 0.97% CFR) and Eastern Africa, where no deaths were reported. These findings are consistent with the well-established higher virulence of Congo Basin (Clade I) viruses compared to West African (Clade II) strains [5,66].

The highest pooled severity rates were observed in Congo (90%) and the Central African Republic (60.9%). This finding is consistent with previous studies reporting high case fatality rates (CFR) of up to 11% in these countries [2,10]. In contrast, outbreaks in Nigeria (2017–2019), which were associated with the less virulent Clade II, exhibited lower CFRs of approximately 2.8% [59]. Collectively, these distinct regional patterns reflect underlying differences in the circulating viral lineage, frequency of zoonotic exposure, and healthcare infrastructure. Central Africa, in particular, continues to experience repeated zoonotic spillovers of more virulent clades coupled with limited diagnostic capacity [67,68].

### Viral clade–specific outcomes

Furthermore, our subgroup analysis reinforces the clade-specific differences in mpox virulence. Infections with Clades Ia and Ib were associated with a higher severity rate (∼30%) and measurable mortality. In contrast, Clade II infections were predominantly less severe, with a 0% severity rate and a lower CFR, though estimates for the latter are based on small sample sizes. These clinical trends are supported by molecular studies indicating that Clade I isolates cause higher viremia, broader tissue dissemination, and more efficient human-to-human transmission [5,69,70].

Our findings align with recent genomic studies documenting an evolutionary transition toward strains with reduced virulence. The emergence of Clade Ib, characterized by mutations that attenuate disease severity while enhancing transmissibility, provides a biological explanation for the observed global decline in severe disease [71]. Consequently, while the global mpox burden is decreasing due to this viral attenuation, severe outcomes remain concentrated in regions where the historically more virulent Clade I persists.

### Study setting & design differences

Both study setting and design significantly influenced the estimates of mpox outcomes. Community-based studies reported a significantly higher case fatality rate (6.11%) than hospital-based studies (1.77%) (*p* < 0.001). This pattern is likely because community-based surveillance captures a broader spectrum of disease, including more severe and late-presenting cases that may not reach healthcare facilities. Similarly, cross-sectional studies, which constituted the majority of the included evidence, yielded higher estimates for both severity (45.6%) and CFR (4.5%) compared to cohort or case series designs. These observations are consistent with earlier meta-analyses that also noted an overrepresentation of severe outcomes in community-based designs [67,72]. Our meta-regression confirmed these findings, identifying a case series design as a significant predictor of higher mortality (*β* = 1.72; *p* = 0.044) and a hospital setting as a predictor of lower mortality (*β* = −1.69; *p* = 0.007).

### Host-related and clinical determinants

Our analysis confirms that host-related risk factors for severe and fatal mpox align with those established in prior literature. HIV co-infection was strongly associated with severe disease and mortality, consistent with reports from Nigeria and the U.S. describing prolonged illness and increased complications in immunocompromised individuals [14,40]. Pediatric age was another critical risk factor, with children under 10 years, and particularly those under 5, facing the highest risk of severe outcomes and death. This echoes historical data where pediatric case fatality rates (CFR) exceeded 15% [12] and is reinforced by recent DRC surveillance indicating that over 80% of mpox deaths occur in children under 15 years [73].

Furthermore, pregnancy and malnutrition were confirmed as significant determinants of poor prognosis [74]. Finally, the absence of smallpox vaccination-derived immunity remains a key driver of susceptibility. This is well-documented by Bunge et al. (2022), who reported higher incidence and severity in younger, unvaccinated cohorts following the cessation of routine smallpox vaccination in the 1980s [2].

### Public health and global implications

Our analysis indicates that despite recent temporal improvements, mpox remains a major endemic challenge in Africa. The continent’s CFR of approximately 4% is an order of magnitude higher than the global average, highlighting persistent and profound inequities in access to vaccines, antivirals, and diagnostic capabilities. To address this, immediate public health priorities must include: (1) expanding genomic surveillance to track the evolution of Clades I and Ib; (2) strengthening vaccination programs in endemic zones using third-generation vaccines; (3) improving diagnostic and clinical management capacity, particularly in rural communities; (4) implementing targeted interventions to protect high-risk groups (children, pregnant women, and immunocompromised individuals); and (5) sustaining international funding and research collaborations to prevent resurgence as global attention wanes.

## Strengths & limitations

This meta-analysis has several key strengths. It is the most comprehensive synthesis of mpox outcomes in Africa to date, covering five decades (1970–2024) across 11 countries, allowing temporal, regional, and genomic comparisons. The inclusion of subgroup and meta-regression analyses, integration of clade-specific data (Ia, Ib, II), and low-to-moderate bias across most studies strengthen the reliability and validity of the findings. Additionally, linking African data with global mpox trends provides valuable context on shifting disease dynamics. Additionally, the small sample sizes in some subgroup analyses result in wide confidence intervals, therefore limiting the precision of the findings. Study in other language than English and French were not searched.

However, the study also has limitations. High heterogeneity (*I*^*2*^ > 80%), limited genomic data constrain comparability. Some regions remain underrepresented, and variations in study design and diagnostic capacity may have influenced estimates. Despite these limitations, the findings offer a robust, continent-wide understanding of mpox severity and mortality trends.

## Conclusions

In comparison with previous literature, this meta-analysis confirms a declining but persistent burden of severe and fatal mpox in Africa, characterized by temporal, geographic, and clade-specific variation. The shift toward milder disease since 2022 mirrors global viral evolution and improved surveillance, yet Central Africa remains at greatest risk due to ongoing Clade I circulation and structural health inequities. Sustained investments in surveillance, vaccination, and health system capacity are essential to reduce morbidity and mortality from mpox in the years ahead.

## Supporting information

Addition Files 1

## Data Availability

The sources of data supporting this systematic review are available in the reference. All data generated or analyzed during this study are included in this published article and supplemental material.

## Abbreviations

*CFR*: Case Fatality Rate
*CI*: Confidence Interval
*DRC*: Democratic Republic of Congo
*HCW*: Healthcare worker
*HIV/AIDS*: Human Immunodeficiency Virus/Acquired Immunodeficiency Syndrome
*MeSH*: Medical Subject Headings
*PCR*: Polymerase Chain Reaction
*PRISMA*: Preferred Reporting Items for Systematic Reviews and Meta-Analysis

## Declarations

### Author contributions

F.Z.L.C. conceived the original idea of the study. F.Z.L.C. conducted the literature search. F.Z.L.C., J.T.Z., C.A. and C.T.A. selected the studies, extracted the relevant information, and synthesized the data. F.Z.L.C. performed the analyses and wrote the first draft of the manuscript. All authors critically reviewed and revised successive drafts of the manuscript. All authors read and approved the final manuscript.

### Ethical approval statement

Not applicable Consent for publication: Not applicable.

