## Additional Files 3 for "Mpox severity and mortality in Africa: a systematic review and meta-analysis"

**Supplementary Table 1** Characteristics of studies included

| Author | Study year*^1^* | Country | Design | Setting | Participant | Sampling | Risk of bias | Suspected cases (n) | Case fatality rate (%) |
| --- | --- | --- | --- | --- | --- | --- | --- | --- | --- |
| Breman *et al.* [1] | 1970 | Liberia | Cross-sectional | Community | General population | Non-probabilistic | L | 4 | 0,0 |
| Breman *et al*. [1] | 1971 | Nigeria | Cross-sectional | Community | General population | Non-probabilistic | L | 2 | 0,0 |
| Breman *et al.* [1] | 1979 | Democratic Republic of Congo | Cross-sectional | Community | General population | Non-probabilistic | L | 38 | 21,1 |
| Arita *et al.* [2] | 1983 | Multinational | Cross-sectional | Community | General population | Non-probabilistic | L | 155 | 11,0 |
| Durski *et al.* [3] | 1984 | Central African Republic | Cross-sectional | Community | General population | Non-probabilistic | M | 6 | 0,0 |
| Jezek *et al.* [4] | 1985 | Democratic Republic of Congo | Cross-sectional | Community | General population | Non-probabilistic | L | 282 | 9,6 |
| Jezek *et al.* [4] | 1986 | Democratic Republic of Congo | Cross-sectional | Community | General population | Non-probabilistic | L | 276 | 2,2 |
| Durski *et al*. [3] | 1987 | Gabon | Cross-sectional | Community | General population | Non-probabilistic | M | 9 | 22,2 |
| Mwamba *et al.* [5] | 1997 | Democratic Republic of Congo | Cross-sectional | Community | General population | Non-probabilistic | L | 92 | 3,3 |
| Hutin *et al.* [6] | 1997 | Democratic Republic of Congo | Cross-sectional | Community | General population | Non-probabilistic | Low | 88 | 0.0 |
| Aplogan *et al.* [7] | 1997 | Democratic Republic of Congo | Cross-sectional | Community | General population | Non-probabilistic | Low | 419 | 0.0 |
| Durski *et al.* [3] | 2003 | Congo | Cross-sectional | Community | General population | Non-probabilistic | Moderate | 12 | 21.1 |
| Boumandouki *et al.* [8] | 2003 | Congo | Case series | Hospital | General population | Non-probabilistic | Low | 8 | 11.0 |
| Formenty *et al.* [9] | 2005 | Sudan | Cross-sectional | Community | General population | Non-probabilistic | Low | 19 | 0.0 |
| Damon *et al.* [10] | 2005 | Sudan | Case report | Community | General population | Non-probabilistic | Low | 2 | 9.6 |
| Durski *et al.* [3] | 2010 | Central African Republic | Cross-sectional | Community | General population | Non-probabilistic | Moderate | 2 | 2.2 |
| Berthet *et al.* [11] | 2010 | Central African Republic | Case report | Community | General population | Non-probabilistic | Low | 2 | 22.2 |
| Reynolds *et al.* [12] | 2010 | Congo | Case series | Community | General population | Non-probabilistic | Low | 11 | 3.3 |
| Pittman *et al.* [13] | 2011 | Democratic Republic of Congo | Cross-sectional | Hospital | General population | Non-probabilistic | Moderate | 222 | 3.4 |
| Durski *et al.* [3] | 2015 | Central African Republic | Cross-sectional | Community | General population | Non-probabilistic | Moderate | 12 | 1.2 |
| Kalthan *et al.* [14] | 2015 | Central African Republic | Cross-sectional | Community | General population | Non-probabilistic | Low | 12 | 8.3 |
| Kalthan *et al.* [15] | 2016 | Central African Republic | Cross-sectional | Community | General population | Non-probabilistic | Low | 26 | 12.5 |
| Nakoune *et al.* [16] | 2016 | Central African Republic | Cross-sectional | Community | General population and HCWs | Non-probabilistic | Moderate | 15 | 0.0 |
| Durski *et al.* [3] | 2017 | Central African Republic | Cross-sectional | Community | General population | Non-probabilistic | Moderate | 2 | 0.0 |
| Durski *et al.* [3] | 2017 | Central African Republic | Cross-sectional | Community | General population | Non-probabilistic | Moderate | 6 | 0.0 |
| Doshi *et al.* [17] | 2017 | Congo | Case series | Community | General population | Non-probabilistic | Moderate | 88 | 100.0 |
| Durski *et al.* [3] | 2017 | Liberia | Cross-sectional | Community | General population | Non-probabilistic | Moderate | 2 | 9.1 |
| Ogoina *et al.* [18] | 2017 | Nigeria | Cross-sectional | Hospital | General population | Non-probabilistic | Low | 38 | 1.4 |
| Ita *et al.* [19] | 2017 | Nigeria | Cross-sectional | Hospital | General population | Non-probabilistic | Low | 15 | 25.0 |
| Ibegu *et al.* [20] | 2017 | Nigeria | Cross-sectional | Hospital | General population | Non-probabilistic | Moderate | 30 | 25.0 |
| Reynolds *et al.* [21] | 2017 | Sierra Leone | Cross-sectional | Community | General population | Non-probabilistic | Low | 2 | 3.8 |
| Vaughan *et al.* [22] | 2018 | Nigeria | Case report | Hospital | General population | Non-probabilistic | Low | 2 | 13.3 |
| Yinka-ogunleye *et al.* [23] | 2018 | Nigeria | Cross-sectional | Community | General population | Non-probabilistic | Low | 276 | 0.0 |
| Ogoina *et al.* [18] | 2018 | Nigeria | Cross-sectional | Hospital | General population | Non-probabilistic | Low | 51 | 0.0 |
| Echekwube *et al.* [24] | 2018 | Nigeria | Case series | Hospital | General population | Non-probabilistic | Low | 4 | 6.8 |
| Ngbolua *et al.* [25] | 2019 | Democratic Republic of Congo | Cross-sectional | Hospital | General population | Non-probabilistic | High | 3 | 0.0 |
| Ogoina *et al.* [26] | 2019 | Nigeria | Cross-sectional | Hospital | General population | Non-probabilistic | Low | 21 | 0.0 |
| Hobson *et al.* [27] | 2021 | Nigeria | Case report | Hospital | General population | Non-probabilistic | Low | 3 | 0.0 |
| Amao *et al.* [28] | 2021 | Nigeria | Cross-sectional | Community | General population | Non-probabilistic | Low | 25 | 0.0 |
| Djuicy *et al.* [29] | 2022 | Cameroon | Cross-sectional | Community and hospital | General population | Non-probabilistic | Low | 137 | 0.0 |
| Besombes *et al.* [30] | 2022 | Central African Republic | Cross-sectional | Community | General population | Non-probabilistic | Low | 11 | 0.0 |
| Yadouleton *et al.* [31] | 2022 | Benin | Case series | Community | General population | Non-probabilistic | Low | 12 | 2.5 |
| Kibungu *et al.* [32] | 2023 | Democratic Republic of Congo | Case series | Community | General population | Non-probabilistic | Low | 5 | 9.8 |
| Nizigiyimana *et al.* [33] | 2024 | Burundi | Cross-sectional | Community | General population | Non-probabilistic | Low | 607 | 0.0 |
| Brosius *et al.* [34] | 2024 | Democratic Republic of Congo | Cohort study | Hospital | General population | Non-probabilistic | Moderate | 511 | 0.0 |
| Mukadi-Bamuleka *et al.* [35] | 2024 | Democratic Republic of Congo | Cross-sectional | Community | General population | Non-probabilistic | Low | 9 | 0.0 |
| Vakaniaki *et al.* [36] | 2024 | Democratic Republic of Congo | Cross-sectional | Community | General population | Non-probabilistic | Low | 241 | 0.0 |
| *1*: Last year that the study was conducted; | | | | | | | | | |

**Study period**

**Event rate (%)**

**Case fatality rate (%)**

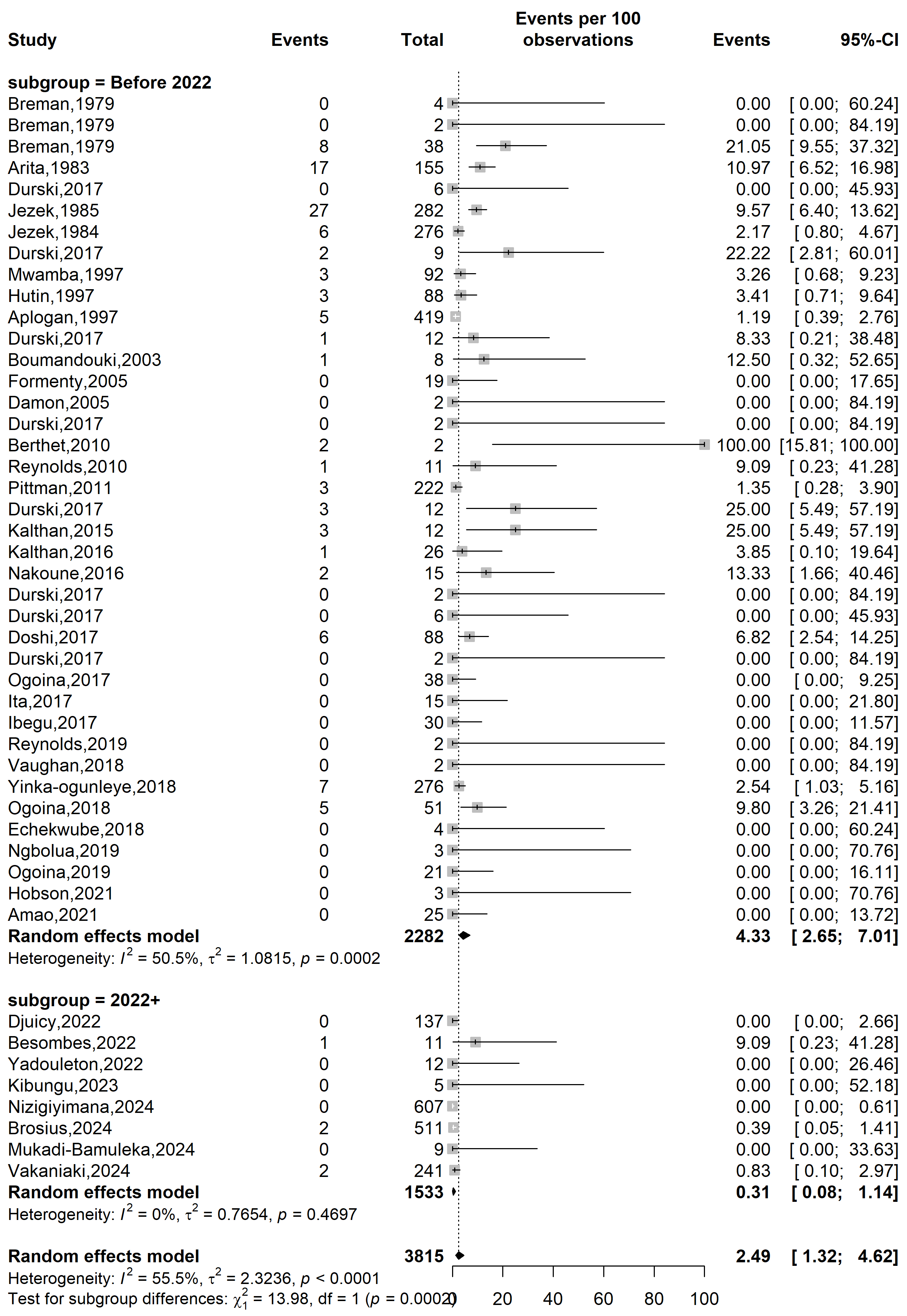

**Supplementary Fig. 1** Case fatality rate among suspected mpox case in Africa by study periods

**Country**

**Event rate (%)**

**Case fatality rate (%)**

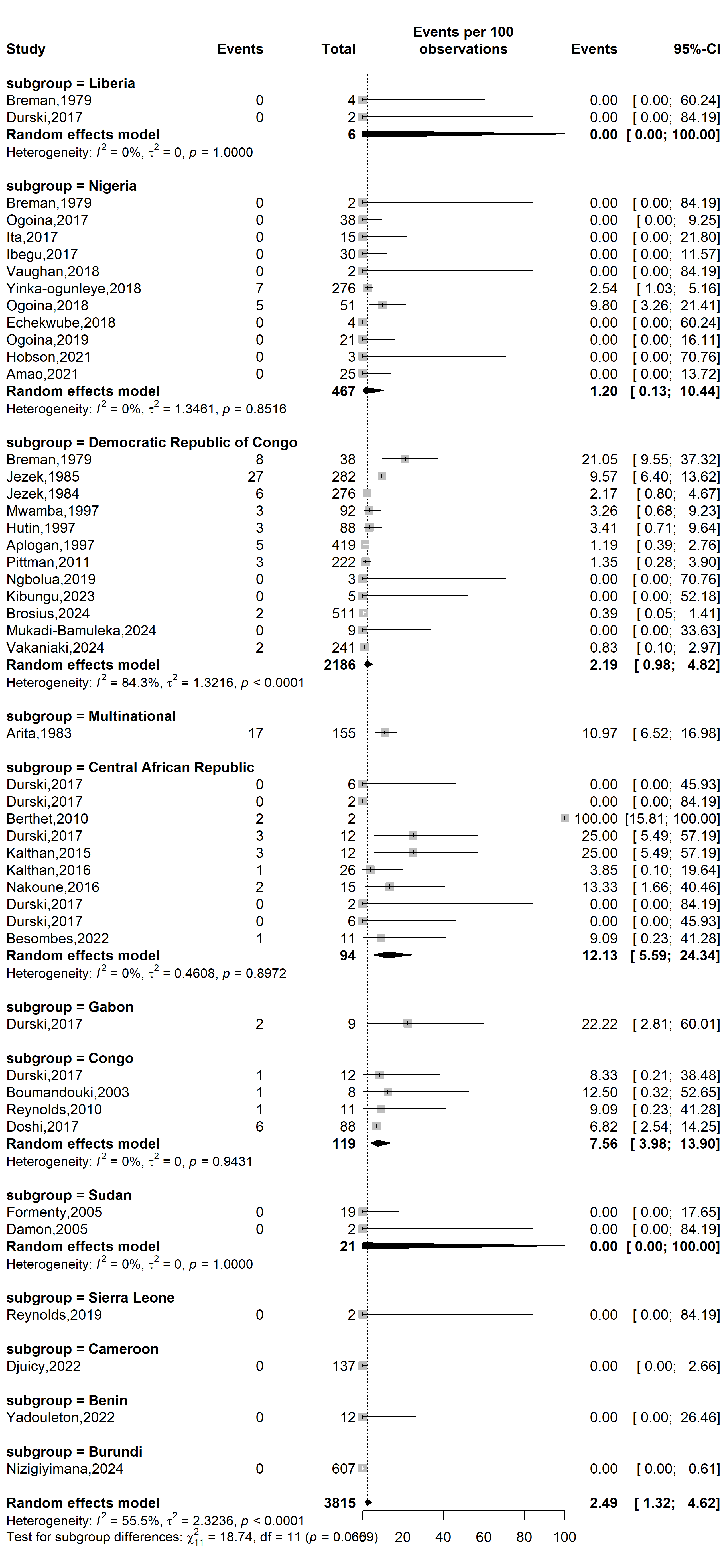

**Supplementary Fig. 2** Case fatality rate among suspected mpox case across African countries

**Study setting**

**Event rate (%)**

**Case fatality rate (%)**

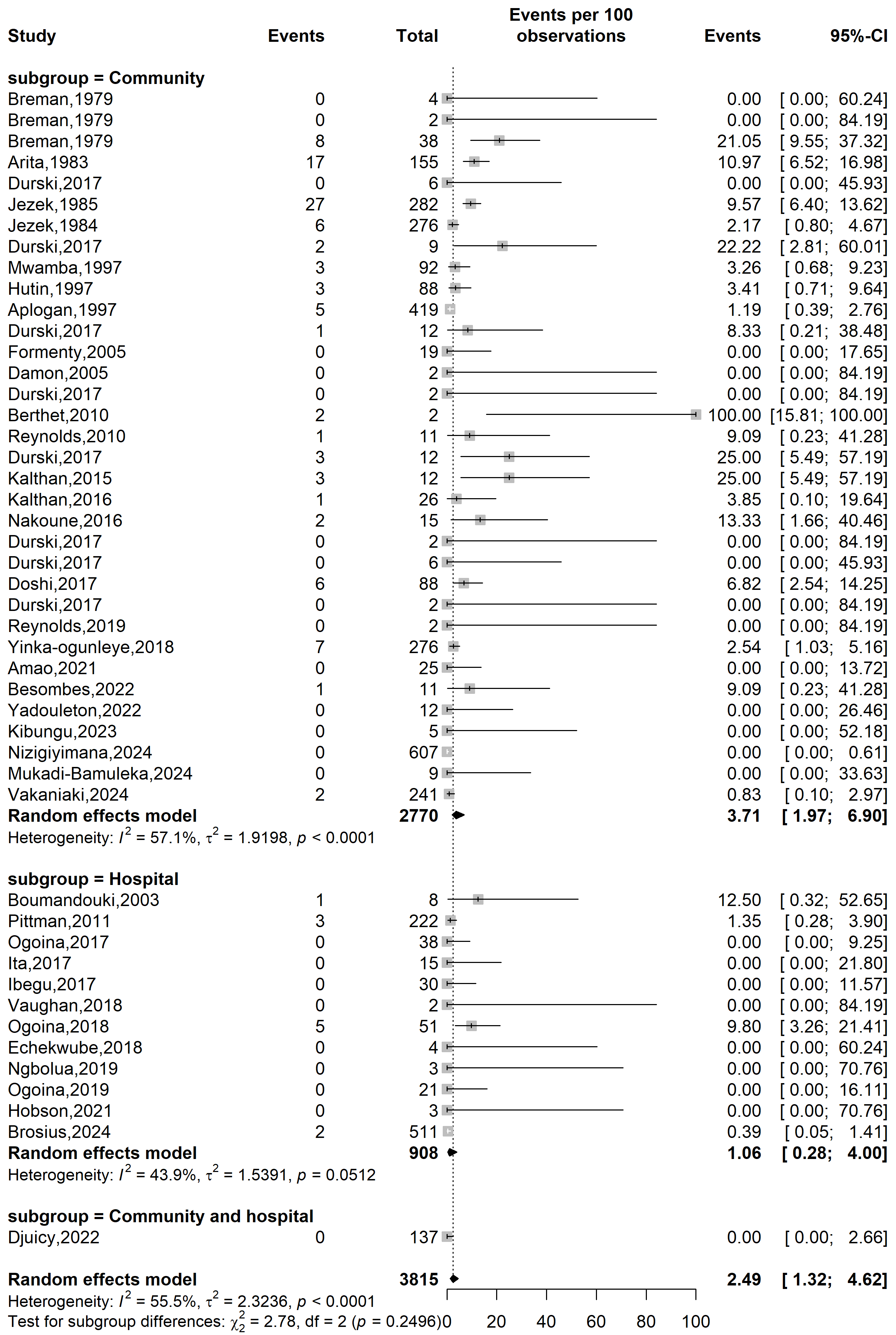

**Supplementary Fig. 3** Case fatality rate among suspected mpox case in Africa by study setting

**Study design**

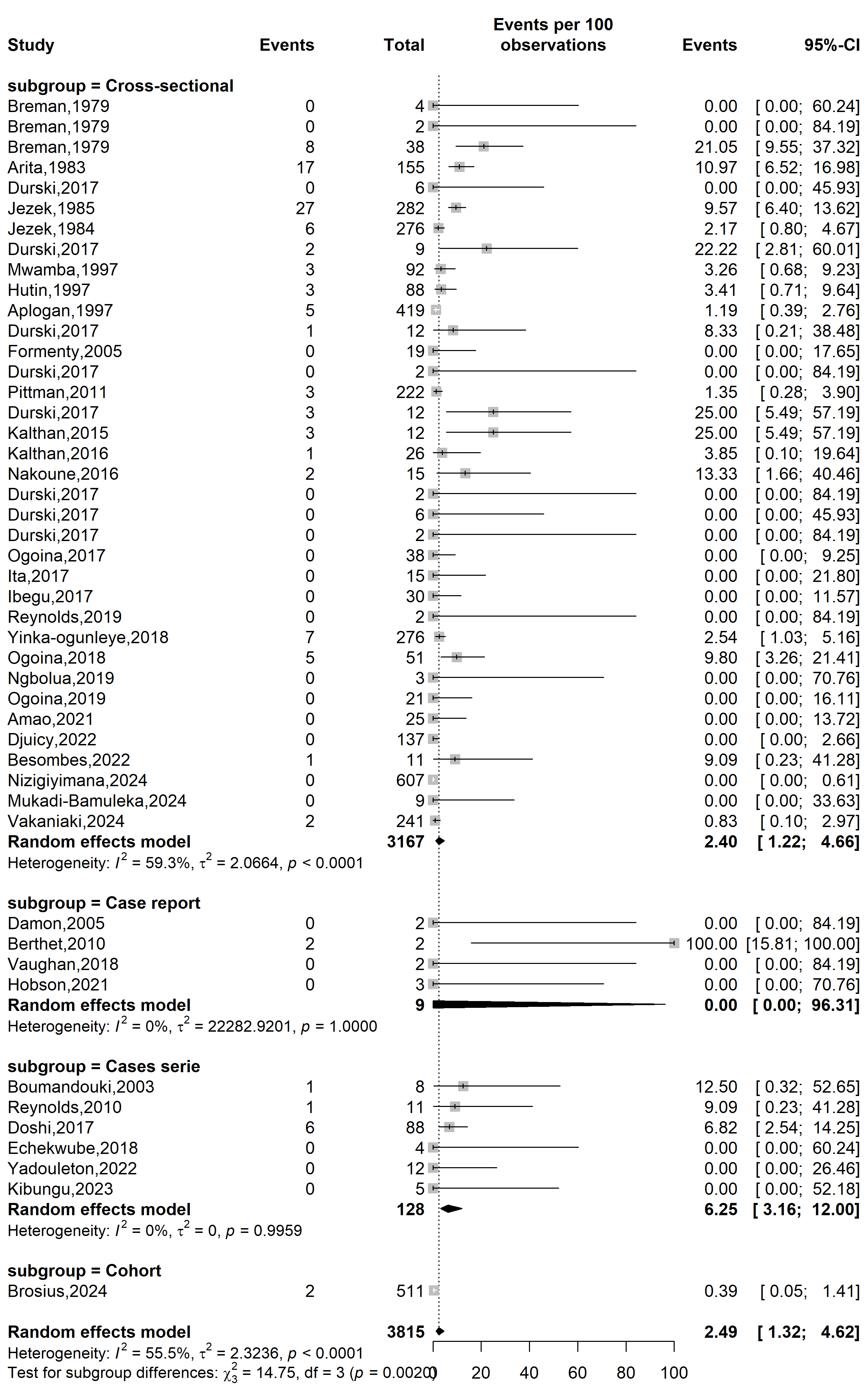

**Case fatality rate (%)**

**Event rate (%)**

**Supplementary Fig. 4** Case fatality rate among suspected mpox case across African countries

Setting

**WHO Afro region**

**Case fatality rate (%)**

**Event rate (%)**

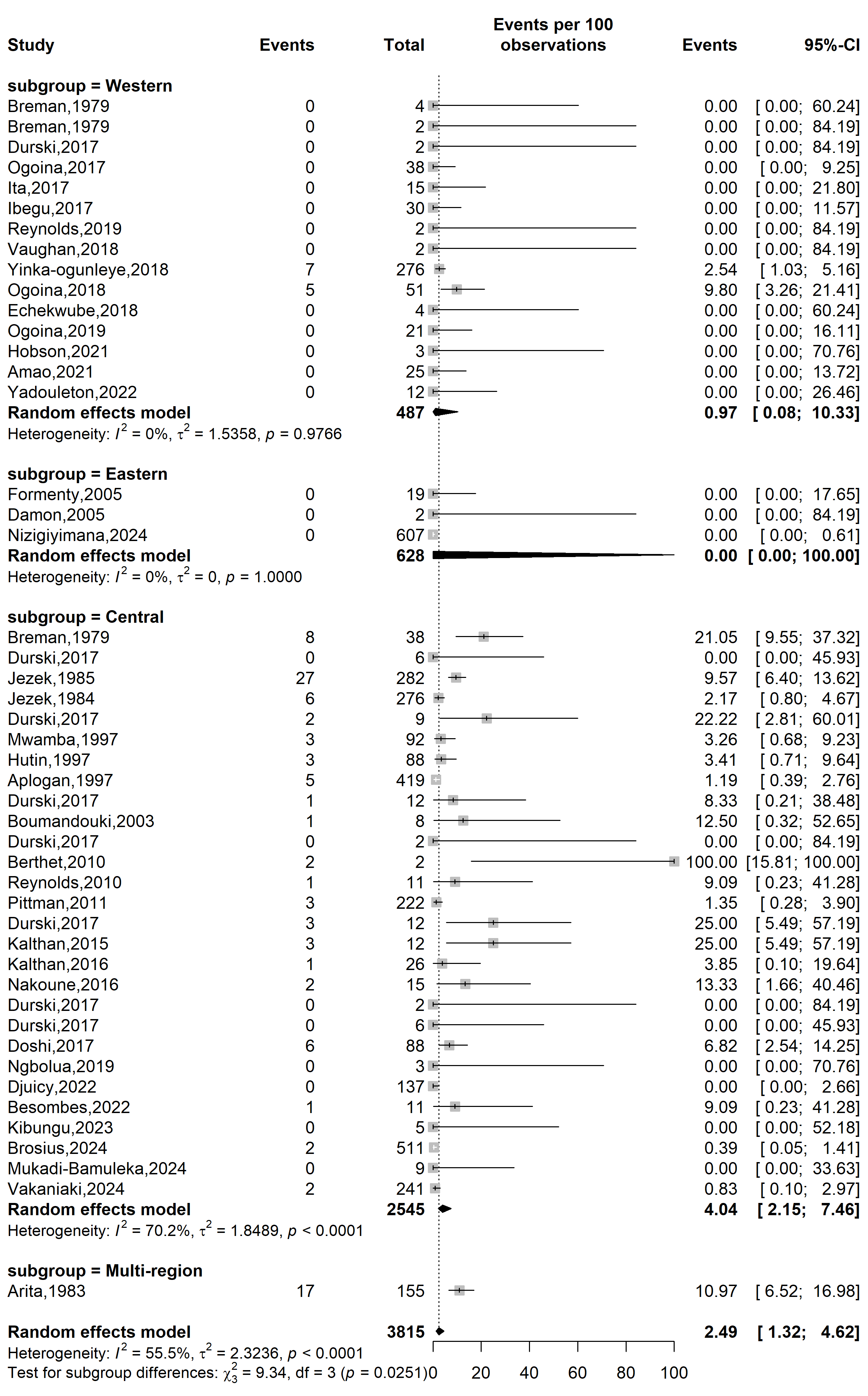

**Supplementary Fig. 5** Case fatality rate among suspected mpox case WHO Afro regions

**Publication bias analysis**

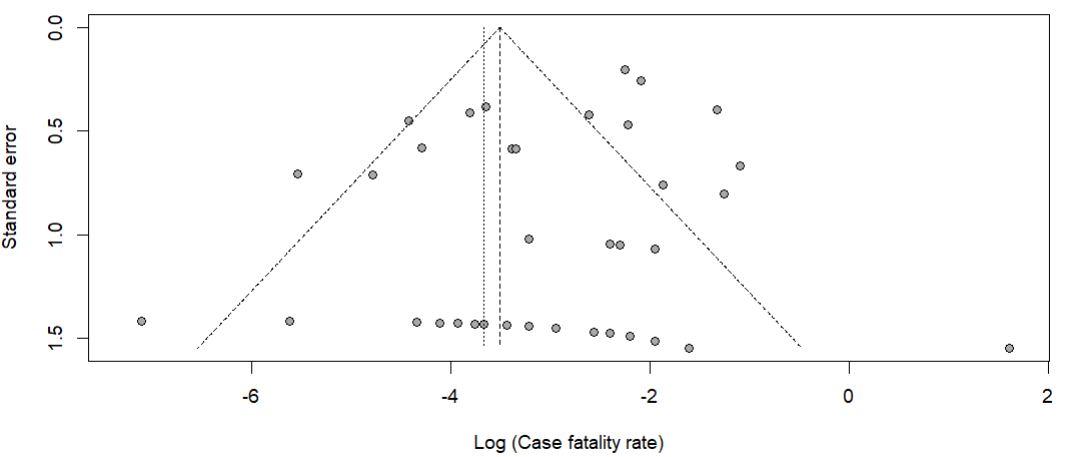

Egger’s test *p*-value = 0.472

Begg’s test *p*-value = 0.019

**Supplementary Fig. 6** Funnel plot displaying the pseudo 95% confidence limits and tests assessing the publication bias of studies included

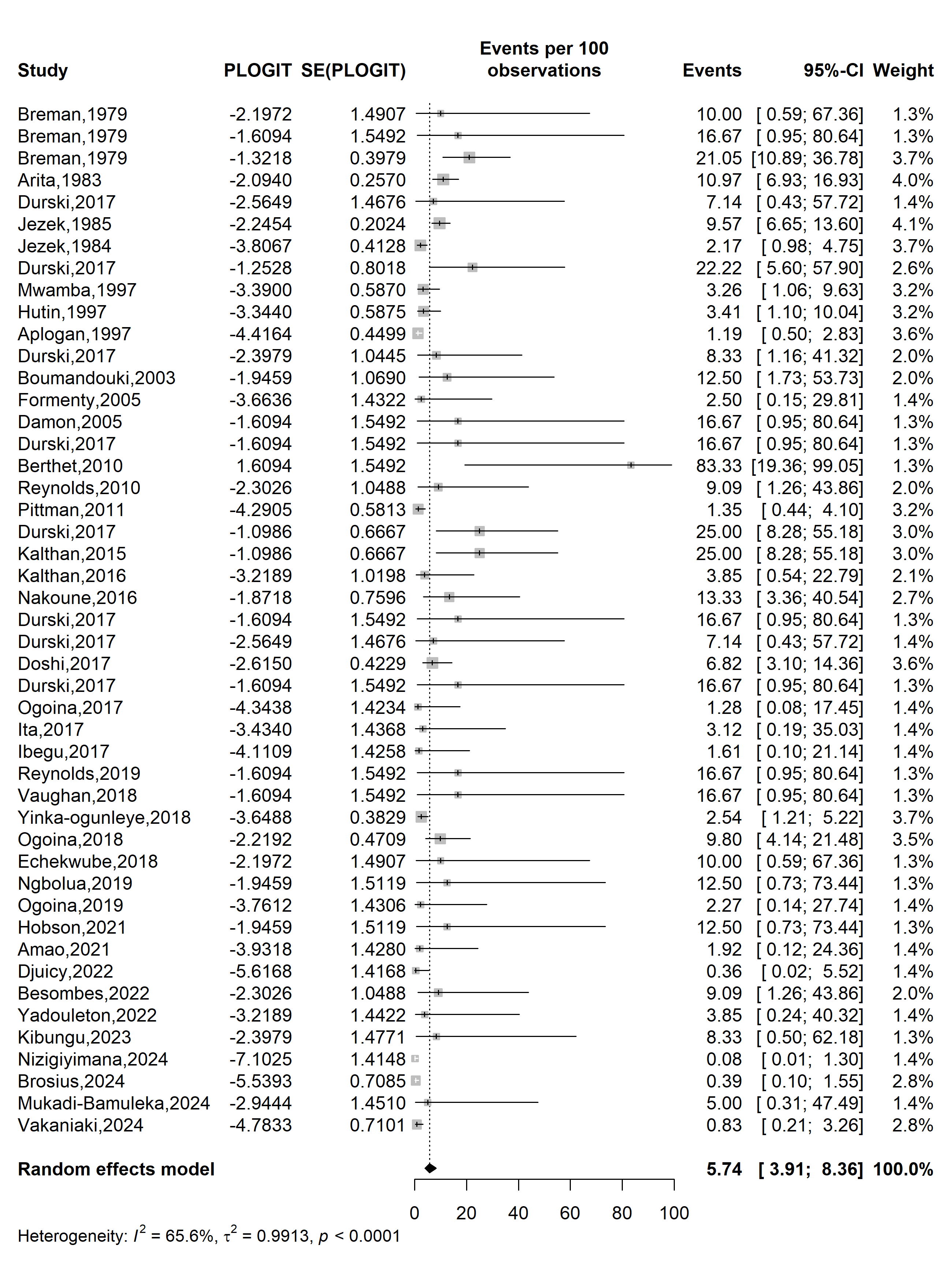

**Event rate (%)**

**Case fatality rate (%)**

**Supplementary Fig. 7** Trim and fill analysis of included studies

**Sensitivity analysis**

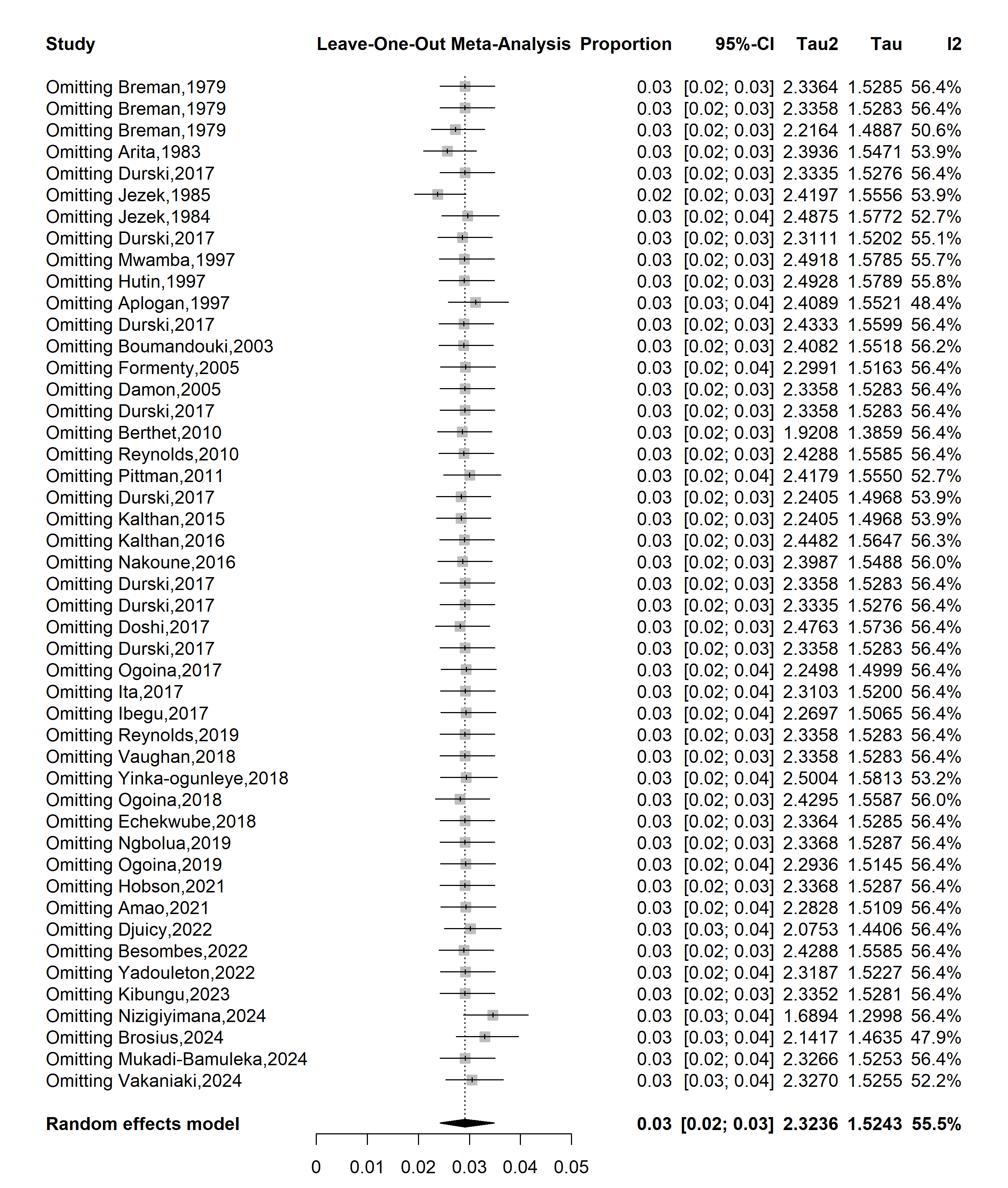

**Supplementary Fig. 8** Sensitivity analysis of the case fatality rate among suspected mpox cases in Africa
