## Supplementary material for "Mpox severity and mortality in Africa: a systematic review and meta-analysis": Addition Files 1

**Supplementary Table 1** Characteristics of studies included

| Author | Study year*^1^* | Country | Design | Setting | Participant | Sampling | Mpox gene sequencing | Risk of bias | Confirmed cases (n) | Severe/grave cases rate (%) |
| --- | --- | --- | --- | --- | --- | --- | --- | --- | --- | --- |
| Breman *et al.* [1] | 1970 | Liberia | Cross-sectional | Community | General population | Non-probabilistic | Not reported | Low | 4 | 0.0 |
| Breman *et al.* [1] | 1971 | Nigeria | Cross-sectional | Community | General population | Non-probabilistic | Not reported | Low | 2 | 50.0 |
| Breman *et al.* [1] | 1979 | Democratic Republic of Congo | Cross-sectional | Community | General population | Non-probabilistic | Not reported | Low | 38 | 57.9 |
| Arita *et al.* [2] | 1983 | Multinational | Cross-sectional | Community | General population | Non-probabilistic | Not reported | Low | 131 | 59.5 |
| Jezek *et al.* [3] | 1985 | Democratic Republic of Congo | Cross-sectional | Community | General population | Non-probabilistic | Not reported | Low | 282 | 39.0 |
| Meyer *et al.* [4] | 2001 | Democratic Republic of Congo | Cross-sectional | Community | General population | Non-probabilistic | Not reported | Low | 16 | 68.8 |
| Boumandouki *et al.* [5] | 2003 | Congo | Case series | Hospital | General population | Non-probabilistic | Not reported | Low | 3 | 100.0 |
| Formenty *et al.* [5] | 2005 | Sudan | Cross-sectional | Community | General population | Non-probabilistic | Clade Ia | Low | 10 | 30.0 |
| Damon *et al.* [6] | 2005 | Sudan | Case report | Community | General population | Non-probabilistic | Clade Ia | Low | 2 | 50.0 |
| Hoff *et al.* [7] | 2007 | Democratic Republic of Congo | Cross-sectional | Community | General population | Non-probabilistic | Not reported | Low | 785 | 36.7 |
| Berthet *et al.* [8] | 2010 | Central African Republic | Case report | Community | General population | Non-probabilistic | Clade Ia | Low | 2 | 0.0 |
| Pittman *et al.* [9] | 2011 | Democratic Republic of Congo | Cross-sectional | Hospital | General population | Non-probabilistic | Not reported | Moderate | 216 | 63.9 |
| Hughes *et al.* [10] | 2014 | Democratic Republic of Congo | Cross-sectional | Community | General population | Non-probabilistic | Not reported | Low | 534 | 36.5 |
| Osadebe *et al.* [11] | 2014 | Democratic Republic of Congo | Cross-sectional | Community | General population | Non-probabilistic | Not reported | Low | 333 | 28.5 |
| Kalthan *et al.* [12] | 2015 | Central African Republic | Cross-sectional | Community | General population | Non-probabilistic | Not reported | Low | 12 | 83.3 |
| Whitehouse *et al.* [13] | 2015 | Democratic Republic of Congo | Cross-sectional | Community | General population | Non-probabilistic | Not reported | Low | 1057 | 50.4 |
| Kalthan *et al.* [14] | 2016 | Central African Republic | Cross-sectional | Community | General population | Non-probabilistic | Not reported | Low | 26 | 61.5 |
| Nakoune *et al.* [15] | 2016 | Central African Republic | Cross-sectional | Community | General population and HCWs | Non-probabilistic | Not reported | Moderate | 10 | 70.0 |
| Doshi *et al.* [16] | 2017 | Congo | Case series | Community | General population | Non-probabilistic | Not reported | Moderate | 7 | 85.7 |
| Ogoina *et al.* [17] | 2017 | Nigeria | Cross-sectional | Hospital | General population | Non-probabilistic | Not reported | Low | 18 | 72.2 |
| Vaughan *et al.* [18] | 2018 | Nigeria | Case report | Hospital | General population | Non-probabilistic | Clade II | Low | 2 | 0.0 |
| Ogoina *et al.* [19] | 2018 | Nigeria | Cross-sectional | Hospital | General population | Non-probabilistic | Not reported | Low | 40 | 60.0 |
| Echekwube *et al.* [20] | 2018 | Nigeria | Case series | Hospital | General population | Non-probabilistic | Not reported | Low | 4 | 25.0 |
| Mande *et al.* [21] | 2019 | Democratic Republic of Congo | Cross-sectional | Community | General population | Non-probabilistic | Not reported | Low | 21 | 38.1 |
| Hobson *et al.* [22] | 2021 | Nigeria | Case report | Hospital | General population | Non-probabilistic | Clade II | Low | 3 | 0.0 |
| Amao *et al.* [23] | 2021 | Nigeria | Cross-sectional | Community | General population | Non-probabilistic | Not reported | Low | 3 | 0.0 |
| Djuicy *et al.* [24] | 2022 | Cameroon | Cross-sectional | Community and hospital | General population | Non-probabilistic | Clade I and II | Low | 32 | 37.5 |
| Yadouleton *et al.* [25] | 2022 | Benin | Case series | Community | General population | Non-probabilistic | Clade II | Low | 3 | 0.0 |
| Besombes *et al.* [26] | 2022 | Central African Republic | Cross-sectional | Community | General population | Non-probabilistic | Clade Ia | Low | 14 | 42.9 |
| Kibungu *et al.* [27] | 2023 | Democratic Republic of Congo | Case series | Community | General population | Non-probabilistic | Clade Ia | Low | 5 | 0.0 |
| Mmerem *et al.* [28] | 2023 | Nigeria | Cohort study | Hospital | General population | Non-probabilistic | Not reported | Low | 56 | 66.1 |
| Nizigiyimana *et al.* [29] | 2024 | Burundi | Cross-sectional | Community | General population | Non-probabilistic | Clade Ib | Low | 154 | 29.2 |
| Brosius *et al.* [30] | 2024 | Democratic Republic of Congo | Cohort study | Hospital | General population | Non-probabilistic | Clade Ib | Moderate | 431 | 24.8 |
| Mukadi-Bamuleka *et al.* [31] | 2024 | Democratic Republic of Congo | Cross-sectional | Community | General population | Non-probabilistic | Clade Ib | Low | 9 | 66.7 |
| Kombozi *et al.* [32] | 2024 | Democratic Republic of Congo | Cross-sectional | Community | General population | Probabilistic | Not reported | Low | 230 | 44.3 |
| Vakaniaki *et al.* [33] | 2024 | Democratic Republic of Congo | Cross-sectional | Community | General population | Non-probabilistic | Clade Ib | Low | 108 | 9.3 |
| Malembi *et al.* [34] | 2024 | Democratic Republic of Congo | Cross-sectional | Community | General population | Non-probabilistic | Clade Ib | Low | 814 | 48.9 |
| *1*: Last year of study implementation; | | | | | | | | | | |

**Study year**

**Event rate (%)**

**Severity rate (%)**

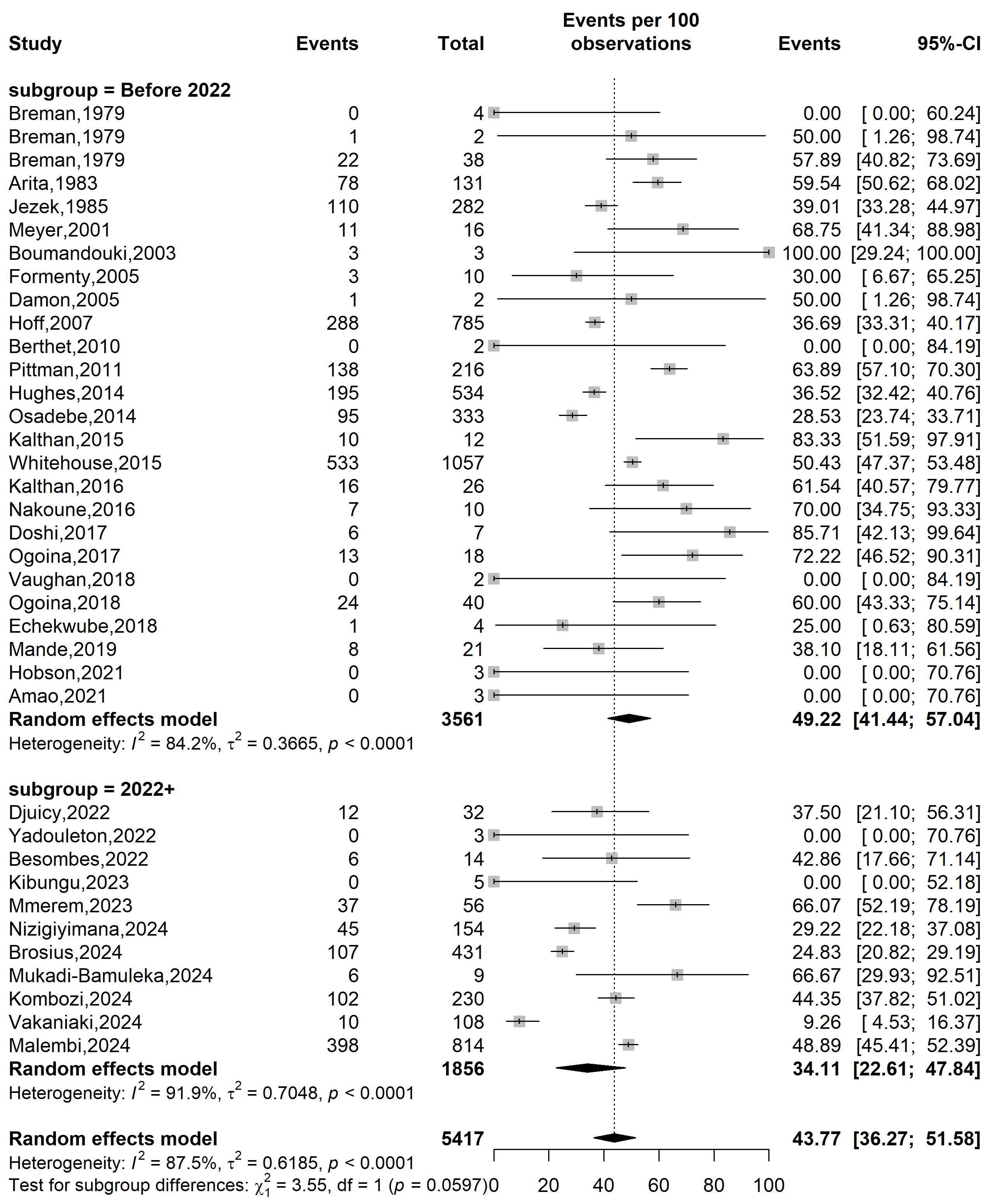

**Supplementary Fig. 1** Mpox disease severity rate in Africa by study periods

**Country**

**Event rate (%)**

**Severity rate (%)**

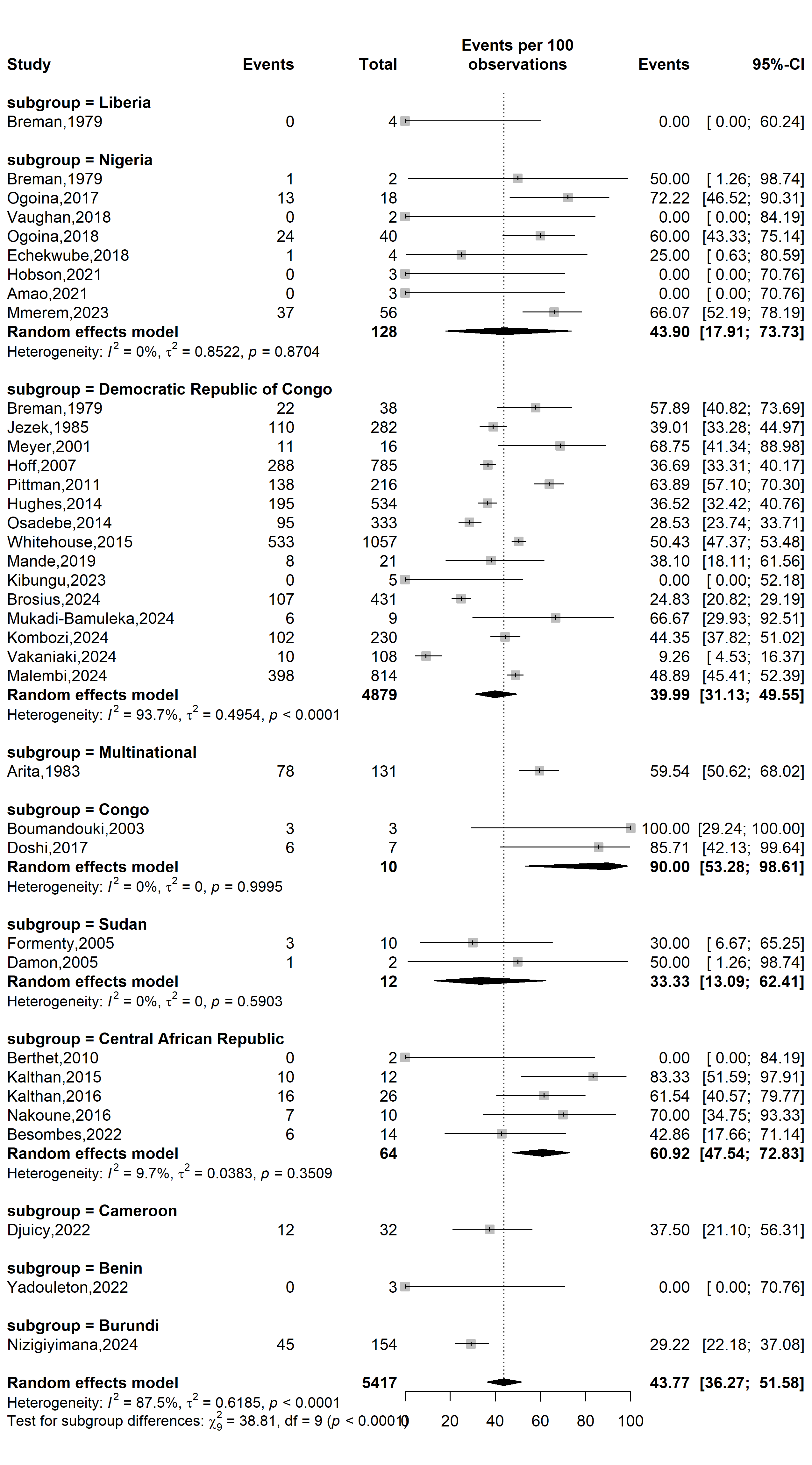

**Supplementary Fig. 2** Mpox disease severity rate in Africa by countries

**WHO Afro region**

**Event rate (%)**

**Severity rate (%)**

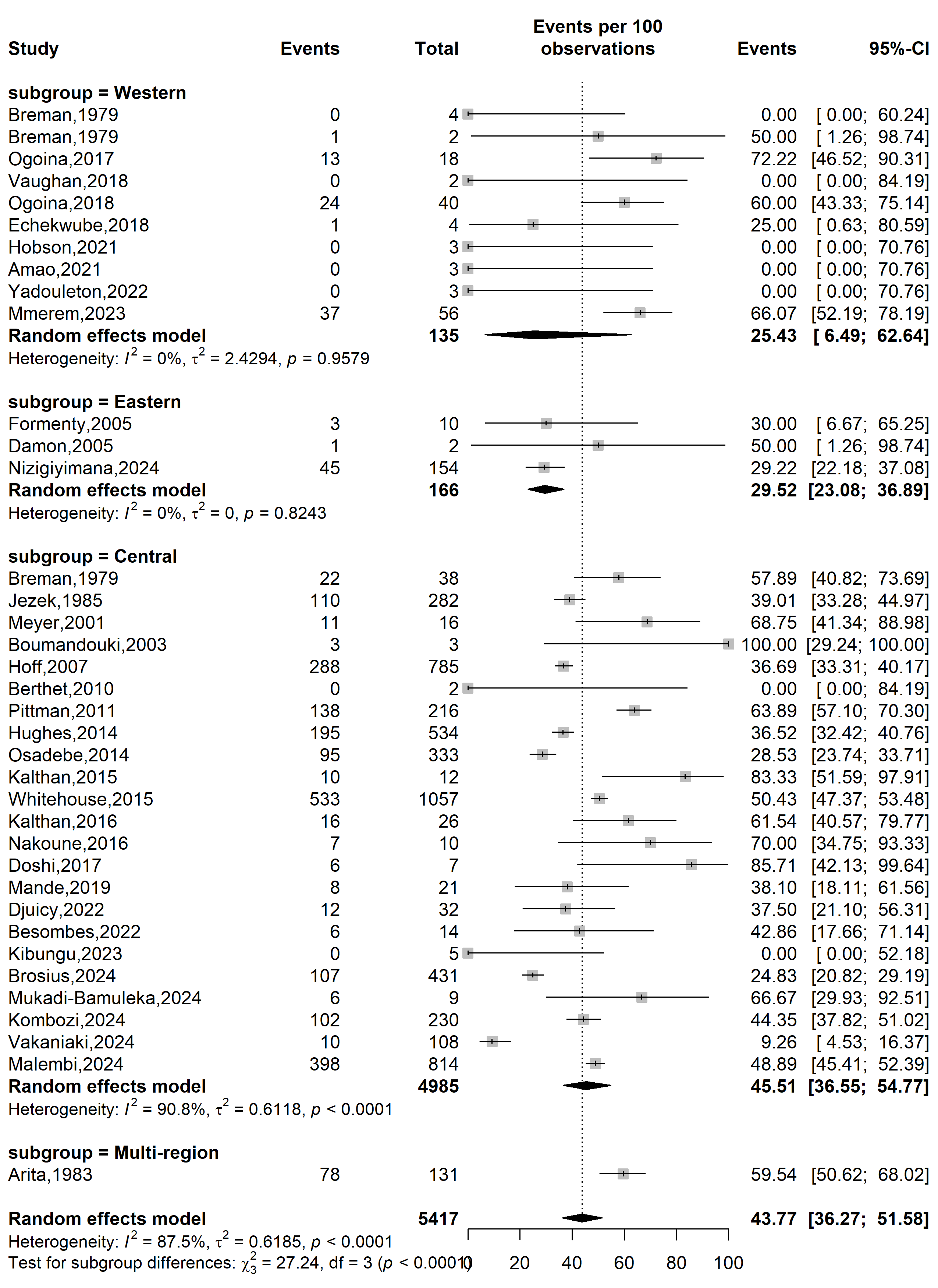

**Supplementary Fig. 3** Mpox disease severity rate in Africa by WHO Afro regions

**Study design**

**Event rate (%)**

**Severity rate (%)**

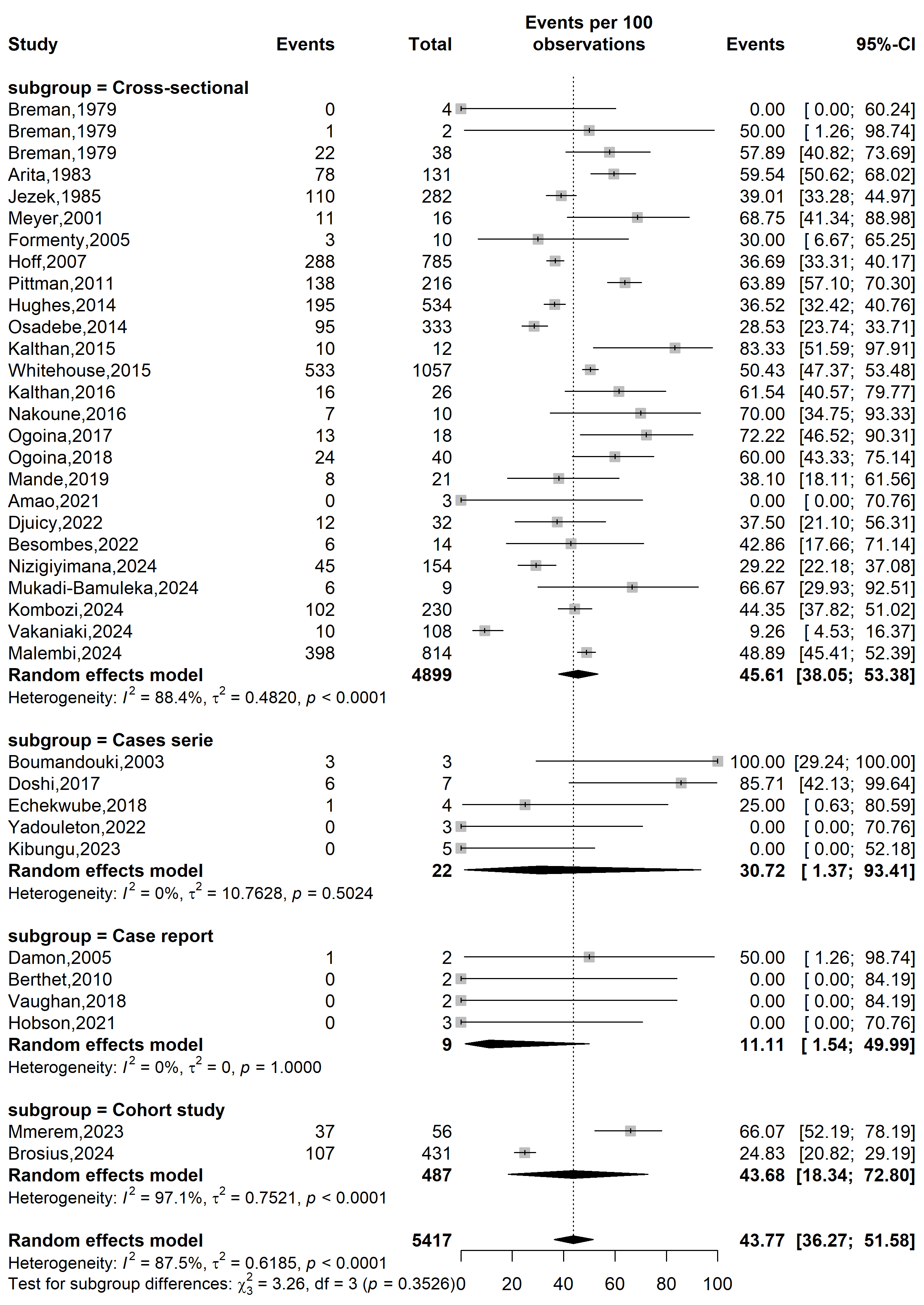

**Supplementary Fig. 4** Mpox severity rate in Africa by type of study designs

**Study setting**

**Event rate (%)**

**Severity rate (%)**

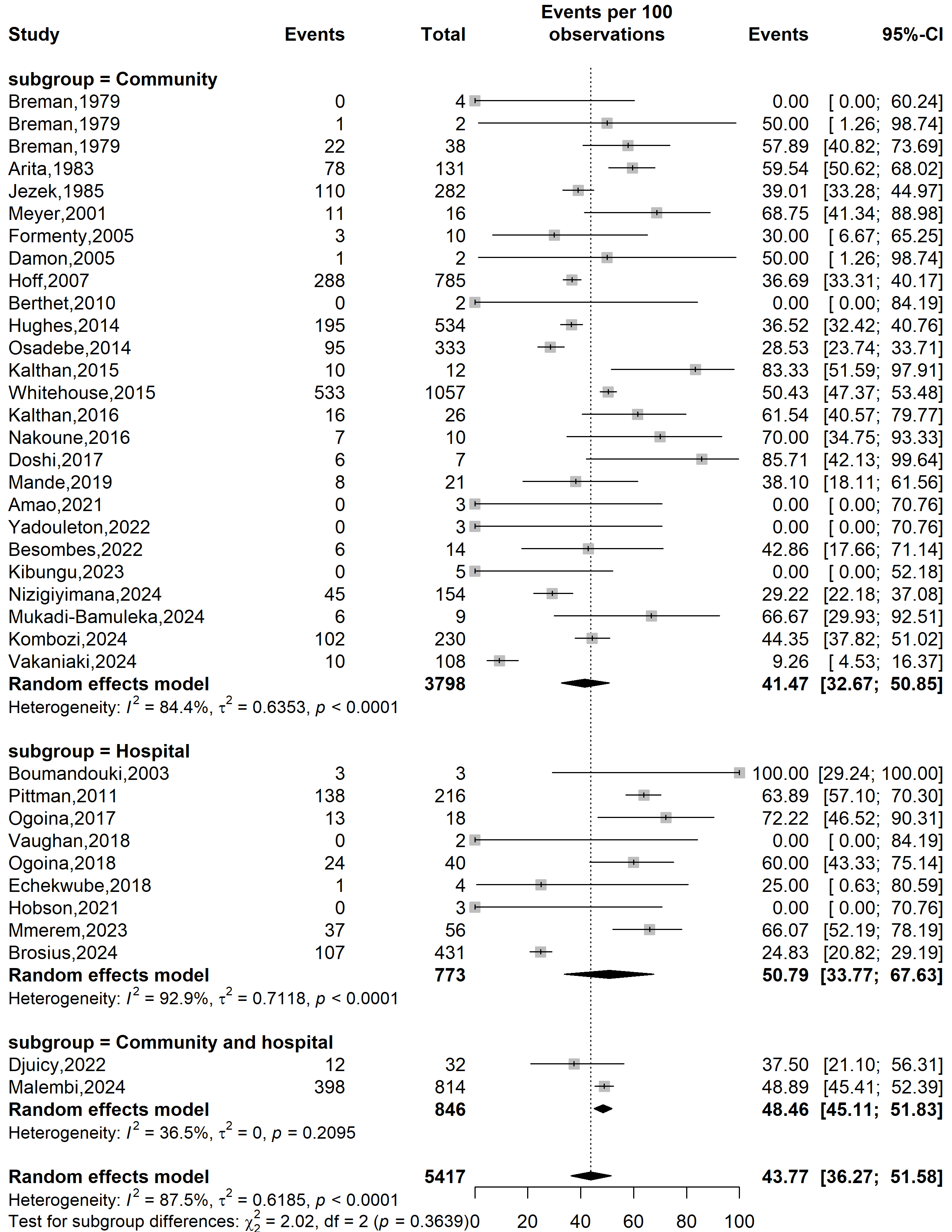

**Supplementary Fig. 5** Mpox severity rate in Africa by type of study settings

**Mpox clade stratification**

**Event rate (%)**

**Severity rate (%)**

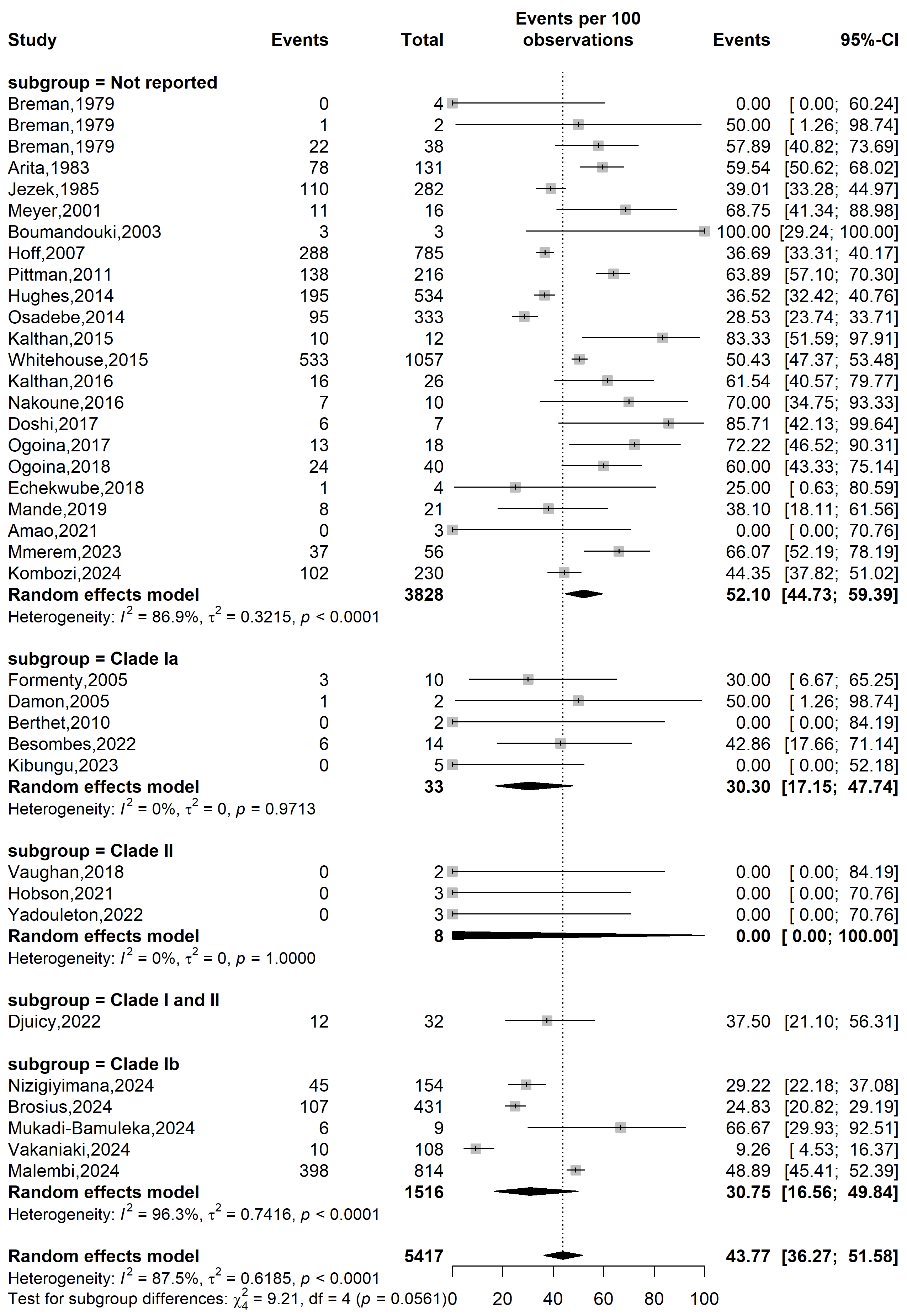

**Supplementary Fig. 6** Mpox severity rate in Africa by clade stratifications

**Publication bias assessment**

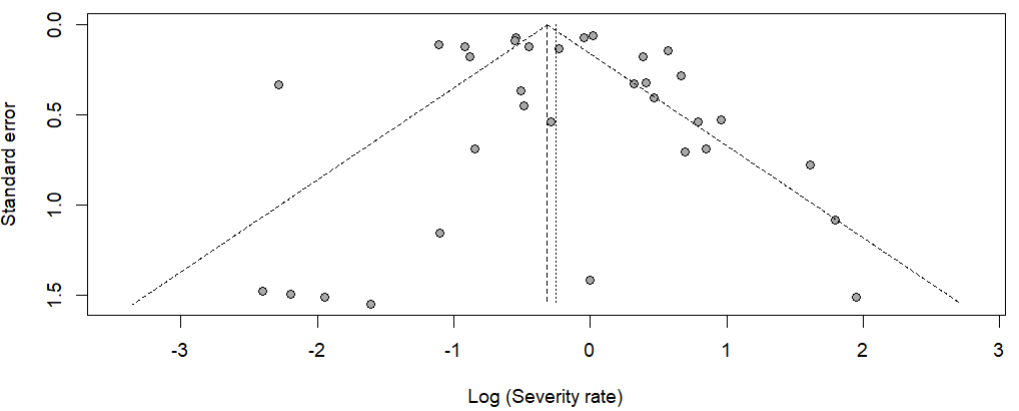

Egger’s test *p*-value = 0.891

Begg’s test *p*-value = 0.326

**Supplementary Fig. 7** Funnel plot displaying the pseudo 95% confidence limits and tests assessing the publication bias of studies included

**Sensitivity analysis**

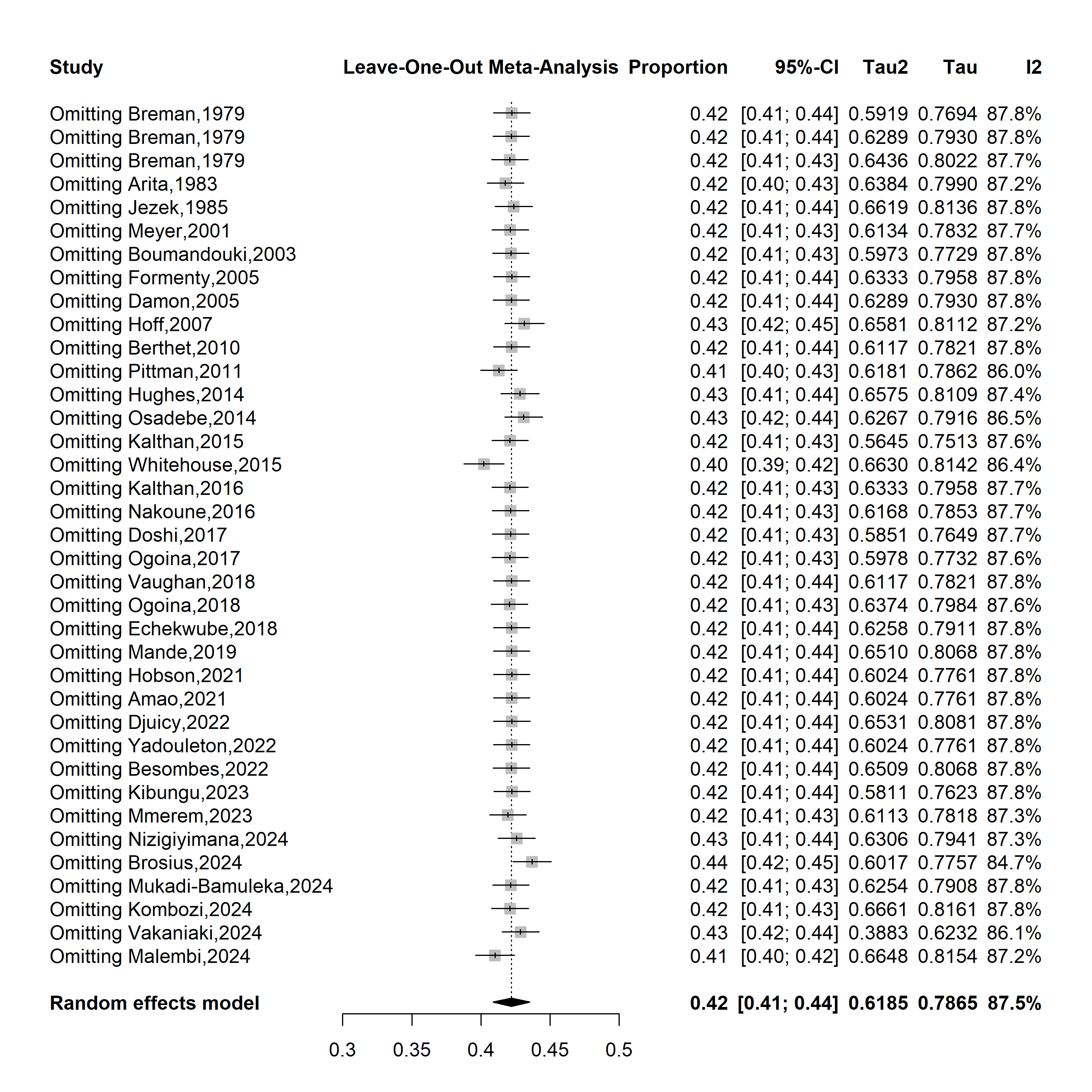

**Supplementary Fig. 8** Sensitivity analysis of the mpox disease severity rate in Africa
